# Beyond Discrete Classifications: A Computational Approach to the Continuum of Cognition and Behavior in Children

**DOI:** 10.1101/2025.04.14.25325835

**Authors:** Anthony Gagnon, Virginie Gillet, Anne-Sandrine Desautels, Jean-François Lepage, Andrea A. Baccarelli, Jonathan Posner, Maxime Descoteaux, Marie A. Brunet, Larissa Takser

## Abstract

Psychiatry is undergoing a shift toward precision medicine, demanding personalized approaches that capture the complexity of cognition and behavior. Here, we introduce a novel referential of four robust, replicable, and generalizable cognitive and behavioral profiles. These were derived from the most prominent pediatric cohort (n=10,843) and validated in two independent cohorts (n=195 and n=271). We demonstrate the profiles’ longitudinal stability and consistency with clinical diagnoses while exposing critical discrepancies across parent-reported, youth-reported, and expert-derived diagnoses. Beyond validation, we showcase the real-world utility of our approach by linking profiles to environmental factors, revealing associations between parental influences and youths’ cognition and behavior. Our fuzzy profiling framework moves beyond discrete classification, offering a powerful tool to refine psychiatric evaluation and intervention. We provide an open-source framework, enabling researchers and clinicians to fast-track implementation and foster a data-driven, domain-based approach to diagnosis. Our findings advocate for broadening the scope of psychiatric assessment.

## Main

The recent years in clinical psychiatry have seen a fueled debate surrounding the current classification systems and their inadequacy to capture the complex nature of psychiatric disorders^1^. One of the core issues with current classification systems, such as the Diagnostic and Statistical Manual of Mental Disorders fifth edition (DSM-V), is the inability to handle the known heterogeneity within a single clinical population as well as the overlap with other populations^2^. New initiatives, such as the Research Domain Criteria (RDoC)^3^ and Hierarchical Taxonomy of Psychopathology (HiTOP)^4^, have proposed frameworks shifting from the traditional categorical view to a dimensional and domain-specific approach. Those frameworks rely on fundamental knowledge in multiple areas of basic science or on data-driven methods to generate empirical domains. Even if they are not widely used in clinical practice, they represent the first steps toward precision medicine in psychiatry and highlight the need to move away from categorical frameworks.

Psychiatric disorders are known to be much more complex than originating from a single cause or defective mechanism, urging clinicians to include environmental, lifestyle, and biological factors within the diagnosis process^5^. However, the influence of environmental factors on psychopathology (e.g., psychiatric disorders) is not fully understood primarily due to the inability to capture the complexity of the symptoms presentation^6^. A plethora of studies have shown relationships between environmental factors, such as supportive social relations, the experience of discrimination, and general exposure to adversity with global mental health/psychopathology^7,8^. In addition, developmental trajectories can be affected by exposure to environmental factors, especially in children, where most disorders onset^9^, remaining an essential aspect of the transition toward personalized medicine. Previous studies support those effects on developmental trajectories; leveraging the Adolescent Brain Cognitive Development (ABCD) cohort^10^, researchers found that familial and community factors, such as family income, marital status, and school environment, significantly predicted cognitive outcomes in pre-adolescents^11^. In the same study, behavioral problems in pre-adolescents were mainly predicted by family conflict, severe financial difficulty, sleep problems, and maternal medical conditions^11^. Additional studies in the same population highlighted an intrinsic relationship between the structure of cognitive abilities and behavioral manifestations, suggesting that worse cognitive abilities are associated with higher behavioral symptoms^12,13^. However, those studies were performed in a single cohort, therefore not extending the generalizability of those results to different populations. As in many fields, generalizable results are hard to achieve and require collecting data from multiple independent populations. Modeling cognition and behavior while retaining the ability to compare and generalize results between populations is still an active open question in modern psychiatry, particularly considering the transition toward personalized medicine. This article tackles this challenge head-on and proposes to model cognition and behavior using fuzzy profiling, embracing the continuum of cognition and behavior found in the general population.

In the past decade, a significant corpus of literature explored extracting profiles from behavioral and psychopathology data in the general population. Indeed, the use of latent class analysis (also named latent profile analysis) has helped uncover profiles showing similar patterns of behavioral symptoms^14^. Although similarities between studies were slight due to different indicator choices or symptoms/disorders of interest, all studies identified relevant subgroups, with the majority reporting the low behavioral symptoms profile as the largest group^14^. One specific study examined the relationship between cognitive measures and behavioral symptoms subgroups, reporting worse working memory, processing speed, and cognitive/intelligence quotient in the internalizing profile and overall worse cognitive performance in the externalizing and dysregulation profiles^15^. While latent profile analysis deals with uncertainty in the form of probabilities (e.g., the likelihood that an event occurs), it merely reflects the confidence in the discrete classification, not the degree of belonging to each profile. This concept makes latent profile analysis suited for scenarios where there should not be an overlap between profiles. However, this is not the case in cognition and behavior; therefore, we must fully embrace the fuzziness of the natural continuum of symptomatology and capabilities found in the general population.

Here, we aimed to create a new referential model that represents the general population’s continuum of cognition and behavior, allowing for direct comparison and generalization between studies. We leveraged data from three independent pediatric cohorts: the ABCD cohort^10^, the Boston Adolescent Neuroimaging of Depression and Anxiety (BANDA) cohort^16^, and the GESTation and Environment (GESTE) cohort^17^. We applied a data-driven fuzzy clustering algorithm within ABCD, the largest pediatric cohort to date, to create our referential cognitive and behavioral profiles. We then showcase the generalizability of our referential profiles by extending them to the two remaining validation cohorts (BANDA and GESTE). Fuzzy logic combined with clustering allows the extraction of patterns, namely profiles, from the data while keeping the natural continuum of cognition and behavior. Unlike probabilistic methods, fuzzy clustering is better suited for overlapping profiles where individuals can exhibit shared characteristics. In classical clustering, two similar subjects are often separated into two different groups by being on each side of the clusters’ boundaries. Therefore, they will be considered entirely differently in subsequent analyses. This separation into groups represents a substantial loss of information compared to continuous methods. Our method enables the retention of this information and the similarity between subjects while extracting meaningful profiles from the data, which is much more suited for real-world scenarios.

Additionally, we demonstrate the stability of our method over a range of developmental periods by independently reproducing the profiles in subsequent follow-ups within the ABCD cohort. Furthermore, by leveraging graph theory concepts, we demonstrate the profiles’ consistency with clinical diagnoses in all cohorts, showing good-to-great consistency. Then, we demonstrate how the profiles can be used to study pressing real-world research questions by investigating the impact of environmental factors on the profiles’ membership values, showing associations between parental factors and youths’ cognition and behavior. The proposed method aligns with the new RDoC ^3^ and HiTOP ^4^ initiatives and supports the need to broaden the scope of the diagnosis process, as it allows the evaluation of symptomatology in an inclusive diagnostic-agnostic manner. This represents a crucial step towards precision medicine since establishing reproducible and stable profiles across developmental periods will enable the study and understanding of clinical trajectories. To facilitate reaching this goal, we provide all the relevant code to reproduce the results in the form of notebooks (https://github.com/Labo-MAB/Gagnon_FuzzyProfiles_2025) and a Python package allowing researchers to use this framework in new populations (https://github.com/gagnonanthony/NeuroStatX).

## Results

From the participants with complete data, 10,843 ABCD participants for the baseline follow-up were included; meanwhile, 195 and 271 participants were retained from the BANDA and GESTE studies, respectively. Demographic information is presented in Table 1. To evaluate the stability of the profiles across developmental stages, 7369 and 2846 participants from ABCD’s 2-year and 4-year follow-ups were included (Supplementary Table 12).

**Table 1.** Demographics Table for all study populations. GED: General Equivalent Diploma. AD: Anxiety Disorder. ADHD: Attention Deficit-Hyperactivity Disorder. CD: Conduct Disorder. DD: Depression Disorder. OCD: Obsessive-Compulsive Disorder. ODD: Oppositional Defiant Disorder. *: Includes all University diplomas (bachelor’s, postgraduates, etc.). ^δ^: Amounts were converted from CAD to USD currency. ^φ^: The mean and std values presented were computed after harmonization. Unavailable data or empty groups are marked with a hyphen.

|  | ABCD |  | BANDA |  | GESTE |  |
| --- | --- | --- | --- | --- | --- | --- |
|  | Male | Female | Male | Female | Male | Female |
| **N (%)** | 5657 (52.17) | 5183 (47.8) | 66 (33.85) | 129 (66.15) | 150 (55.35) | 121 (44.65) |
| **Age, months (std)** | 119.1 (7.52) | 118.82 (7.49) | 181.86 (9.69) | 187.18 (9.73) | 137.54 (11.83) | 136.93 (11.31) |
| **Race/Ethnicity, count (%)** |  |  |  |  |  |  |
| White | 3033 (27.97) | 2657 (24.5) | 49 (25.13) | 90 (46.15) | 123 (45.39) | 98 (36.16) |
| Black or African American | 804 (7.41) | 817 (7.53) | 1 (0.51) | 4 (2.05) | 1 (0.37) | - |
| Hispanic or Latino | 1123 (10.36) | 1037 (9.56) | 5 (2.56) | 9 (4.62) | - | 2 (0.74) |
| Asian | 110 (1.01) | 126 (1.16) | 1 (0.51) | 6 (3.08) | - | - |
| Other | 587 (5.41) | 546 (5.04) | 10 (5.13) | 20 (10.26) | 26 (9.59) | 21 (7.75) |
| **Highest parental education, count (%)** |  |  |  |  |  |  |
| No Highschool | 238 (2.19) | 270 (2.49) | - | - | - | - |
| Highschool, GED, or equivalent | 531 (4.9) | 482 (4.45) | - | - | 7 (2.58) | 6 (2.21) |
| Some college | 1478 (13.63) | 1327 (12.24) | 9 (4.62) | 8 (4.1) | 78 (28.78) | 57 (21.03) |
| Bachelor's Degree | 1458 (13.45) | 1304 (12.03) | 13 (6.67) | 37 (18.97) | 60 (22.14) * | 54 (19.93) * |
| Postgraduate Degree | 1941 (17.9) | 1797 (16.57) | 44 (22.56) | 75 (38.46) |  |  |
| **Familial Income (USD\$), count (%)** | | | | | | |
| < 50 000\$ | 1053 (9.71) | 999 (9.21) | - | - | 32 (11.81) <sup>δ</sup> | 28 (10.33) <sup>δ</sup> |
| 50 000 - 100 000\$ | 1907 (17.59) | 1750 (16.14) | - | - | 59 (21.77) <sup>δ</sup> | 48 (17.71) <sup>δ</sup> |
| > 100 000\$ | 2211 (20.39) | 2002 (18.46) | - | - | 37 (13.65) <sup>δ</sup> | 26 (9.59) <sup>δ</sup> |
| **Psychopathology, count (%)** |  |  |  |  |  |  |
| AD | 563 (5.19) | 535 (4.93) | 32 (16.41) | 36 (18.46) | - | - |
| ADHD | 1103 (10.17) | 556 (5.13) | 20 (10.26) | 24 (12.31) | 37 (13.65) | 15 (5.54) |
| CD | 212 (1.96) | 90 (0.83) | - | - | - | - |
| DD | 23 (0.21) | 16 (0.15) | 19 (9.74) | 61 (31.28) | - | - |
| OCD | 500 (4.61) | 320 (2.95) | 6 (3.08) | 10 (5.13) | - | - |
| ODD | 380 (3.5) | 218 (2.01) | 5 (2.56) | 7 (3.59) | - | - |
| **Cognitive and Behavioral Scores, mean (std) <sup>φ</sup>** |  |  |  |  |  |  |
| Internalization | -0.00 (0.98) | 0.0 (0.98) | -0.03 (0.94) | 0.01 (1.02) | 0.0 (0.97) | -0.0 (1.01) |
| Externalization | 0.01 (1.06) | -0.0 (0.86) | 0.02 (1.13) | -0.0 (0.89) | 0.0 (1.0) | -0.0 (0.94) |
| Stress | 0.0 (1.02) | -0.0 (0.92) | -0.0 (1.02) | 0.0 (0.97) | 0.04 (0.79) | 0.03 (0.75) |
| VA | -0.02 (0.62) | 0.03 (0.61) | -0.03 (0.66) | 0.02 (0.6) | 0.01 (0.66) | -0.02 (0.56) |
| EFPS | -0.02 (0.49) | 0.02 (0.47) | -0.02 (0.5) | 0.01 (0.48) | 0.01 (0.49) | -0.01 (0.45) |
| MEM | -0.02 (0.49) | 0.02 (0.48) | -0.01 (0.43) | 0.01 (0.46) | 0.01 (0.5) | -0.01 (0.48) |

### Cognitive and behavioral fuzzy profiles

For each cohort, we extracted three latent cognitive factors (verbal ability, executive function/processing speed, and memory) using split-sample exploratory and confirmatory factor analysis as proposed in previous studies^12,13^. Additionally, we extracted three behavioral scores (internalization, externalization, and stress) from validated questionnaires. Then, cognitive factors and behavioral scores were residualized for covariates and included in the fuzzy clustering analysis (see Methods, Figure 1, and Table 1).

**Figure 1.**
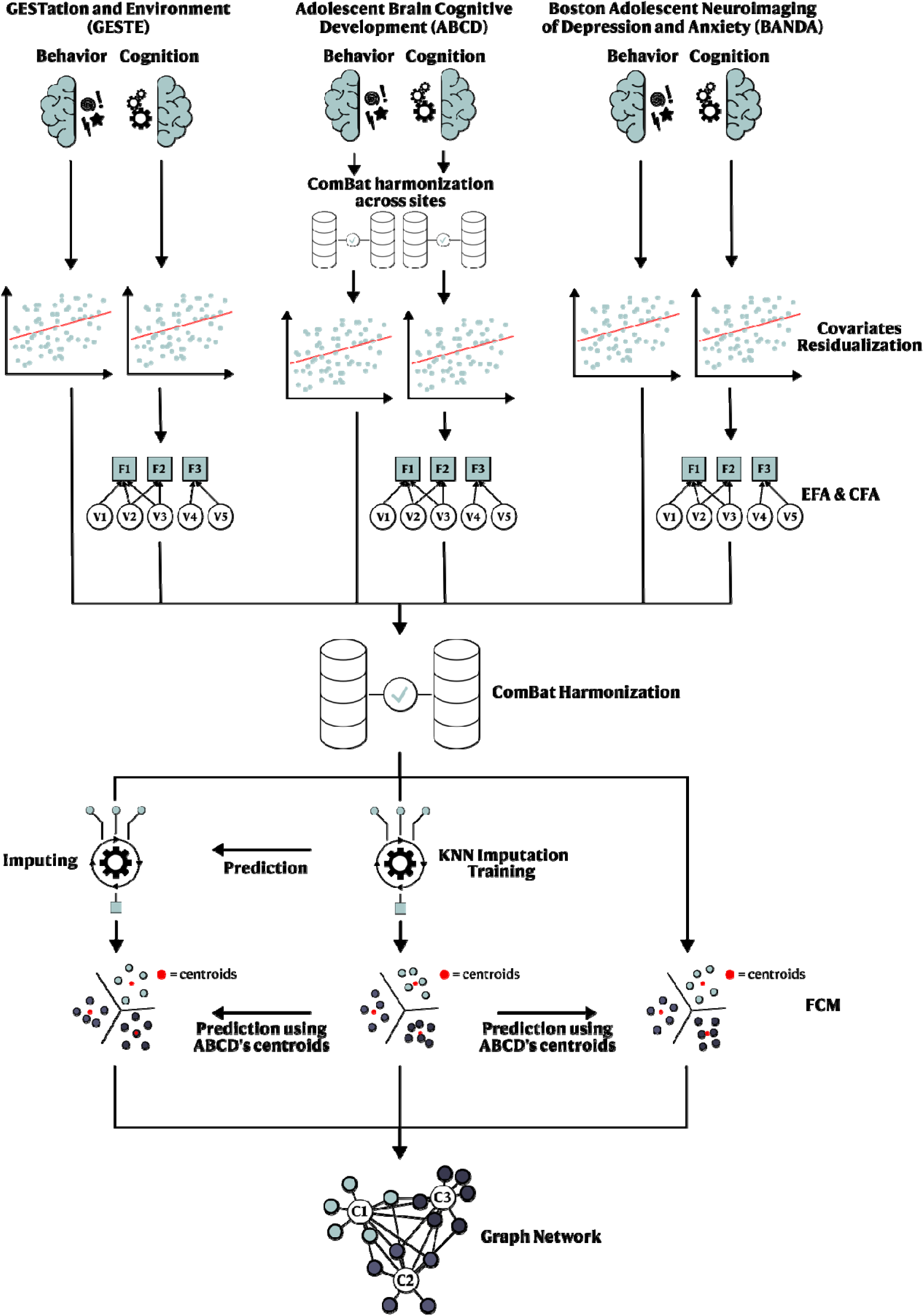
Overview of the statistical pipeline. EFA: Exploratory Factorial Analysis, CFA: Confirmatory Factorial Analysis, KNN: K-nearest Neighbors, FCM: Fuzzy C-Means.

The baseline ABCD study returned an optimal 4-cluster solution, derived using the silhouette score, representing four different behavioral and cognitive profiles (Figure 2, Supplementary Figure 7). Using the participants’ highest membership value as their primary profile, profiles C3 and C4 contain most study participants (n = 4032 and 3730, respectively) and represent low behavioral scores with high (HC/LB) and low (LC/LB) cognitive scores, respectively (Figure 2). Profiles C1 and C2 contain fewer study participants (n = 1896 and 1651, respectively) and show moderate cognitive scores with high stress/internalizing behavior (MC/HSI) and high externalizing behavior scores (MC/HE), respectively (Figure 2). Interestingly, participants with higher behavior scores (all domains included) were only associated with moderate cognitive capabilities, not low or high cognitive capabilities.

**Figure 2.**
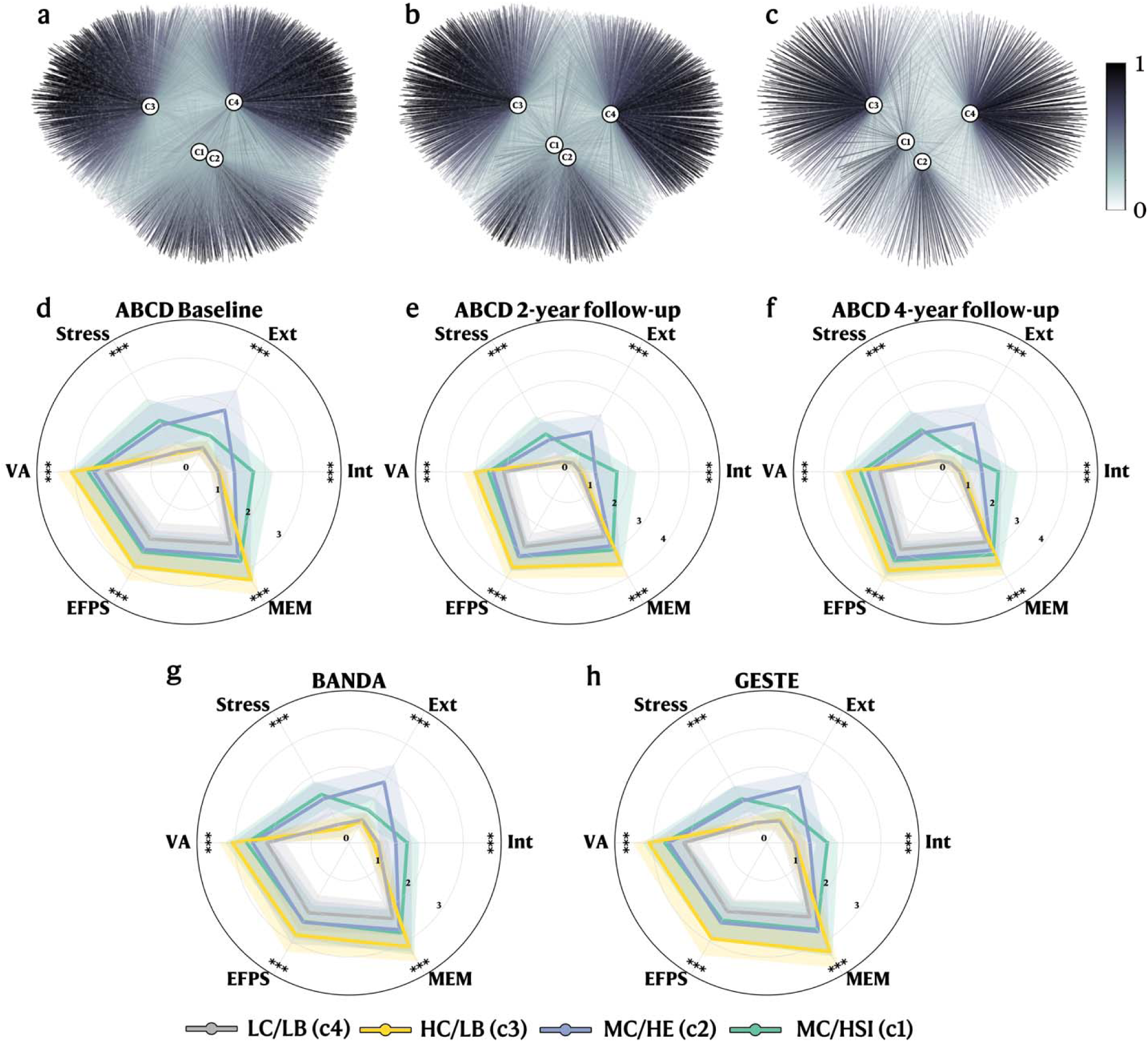
FCM cognitive and behavioral profiles. **a.** Graph network representing the FCM clustering results (on the ABCD baseline data), including the mapped BANDA and GESTE datasets. Grey nodes represent subjects, and edges represent the membership values between a single subject and each cluster’s centroid. **b-c.** Graph networks representing the independent FCM clustering results for ABCD 2-year and 4-year follow-ups. **d-h.** Radar plot of the mean values (with standard deviation) stratified by clusters for each behavioral symptom and cognitive variable for each cohort or follow-up. **d.** ABCD baseline follow-up. **e.** ABCD 2-year follow-up. **f.** ABCD 4-year follow-up. **g.** BANDA. **h.** GESTE. Participants were grouped based on their primary profile (the highest membership value) for the radar plots. ***: *p* < 0.001. Scores are scaled for visualization purposes. VA: Verbal Ability. EFPS: Executive Function and Processing Speed. MEM: Memory. Ext: Externalization. Int: Internalization.

### Longitudinal stability of the profiles

To evaluate the stability across developmental stages of the extracted profiles, we performed additional independent FCM analysis on the 2-year and 4-year follow-ups using identical methods and input variables as the baseline follow-up (see Methods). Both returned nearly identical profiles, with profiles C3 (HC/LB) and C4 (LC/LB) containing most participants (2y: n = 2631 and 2504, 4y: n = 1015 and 1018, respectively) while profiles C1 (MC/HSI) and C2 (MC/HE) contain fewer participants (2y: n = 1130 and 1104, 4y: 436 and 377) (Figure 2). Participants exhibiting higher behavioral scores on all scales were again associated with moderate cognitive abilities.

### Generalizability to external populations

Generalizable results are key to reproducible and sustainable science; therefore, we assess the replicability and robustness of our profiles in two external cohorts. After harmonization, we predicted the membership values for the BANDA and GESTE participants using ABCD clusters’ centroids (see Methods). The prediction process relies on mapping the participants onto the existing ABCD profiles and computing their distance from each centroid. Prediction does not need complete coverage of all profiles and is suitable for studies of smaller sizes (see Methods). Both BANDA profiles (HC/LB: 59, LC/LB: 55, MC/HSI: 47, and MC/HE: 34 participants) and GESTE profiles (HC/LB: 83, LC/LB: 77, MC/HSI: 58, and MC/HE: 53 participants) showed close-to-identical score patterns, highlighting their replicability and robustness across various populations (Figure 2 and Supplementary Figure 8). The difference in means between each profile for each cohort and between cohorts is provided in Supplementary Tables 13, 14, 15, 16, 17, and 18.

### Consistency with clinical diagnoses

Extracting profiles from populational data is relatively easy. However, extracting *meaningful* profiles from populational data is a more challenging task. To validate that the proposed profiles had clinical utility, we evaluated their consistency with existing DSM-V diagnoses. While diagnoses have limitations surrounding heterogeneity and overlap between clinical populations, they represent the current clinical standard that should be reflected within the profiles with expected differences. First, we projected the profiles into a graph network, labeling nodes as participants and edges as membership values; then, we mapped participants with a diagnosis and evaluated their distribution pattern (Figure 3). Considering the highest membership value as the participants’ main profile, we assessed the distribution of anxiety disorder (AD), attention deficit-hyperactivity disorder (ADHD), conduct disorder (CD), depressive disorder (DD), obsessive-compulsive disorder (OCD), and oppositional defiant disorder (ODD) obtained from the parent-administered Kiddie Schedule for Affective Disorders and Schizophrenia for School-Aged Children (KSADS) (ABCD and BANDA) and the medical records (GESTE) (see Methods and Table 1). We found that participants with disorders characterized by externalizing behaviors (ADHD, CD, and ODD) were mainly within the MC/HE profile in the ABCD (39.82%, 74.50%, and 67.39%, respectively) and BANDA studies (ADHD: 38.64%, and ODD: 66.67%) (Figure 3). However, in the GESTE study, in which psychopathology was pulled from medical records, ADHD participants were split between the LC/LB and MC/HE profiles (36.54% and 30.77%, respectively) (Figure 3). Participants with AD or OCD were marginally more present in profile MC/HSI in the ABCD study (37.85% and 42.93%, respectively) (Figure 3). In contrast, participants with a DD diagnosis were equally found within profile MC/HSI and MC/HE in the ABCD and BANDA studies (ABCD: 48.72% and 46.15%, respectively, and BANDA: 38.75% and 30.00%, respectively) (Figure 3). However, AD and OCD in the BANDA study were also found in the MC/HSI profile (35.20% and 43.75%, respectively), whereas OCD was also prevalent in the HC/LB profile (37.50%) (Figure 3). Using the youth-administered KSADS in the ABCD cohort, AD participants were mainly found within the MC/HSI profile (39.06%). In contrast, DD participants were found within the LC/LB, MC/HSI, and MC/HE profiles (36.14%, 25.30%, and 24.10%, respectively) (Supplementary Figure 10).

**Figure 3.**
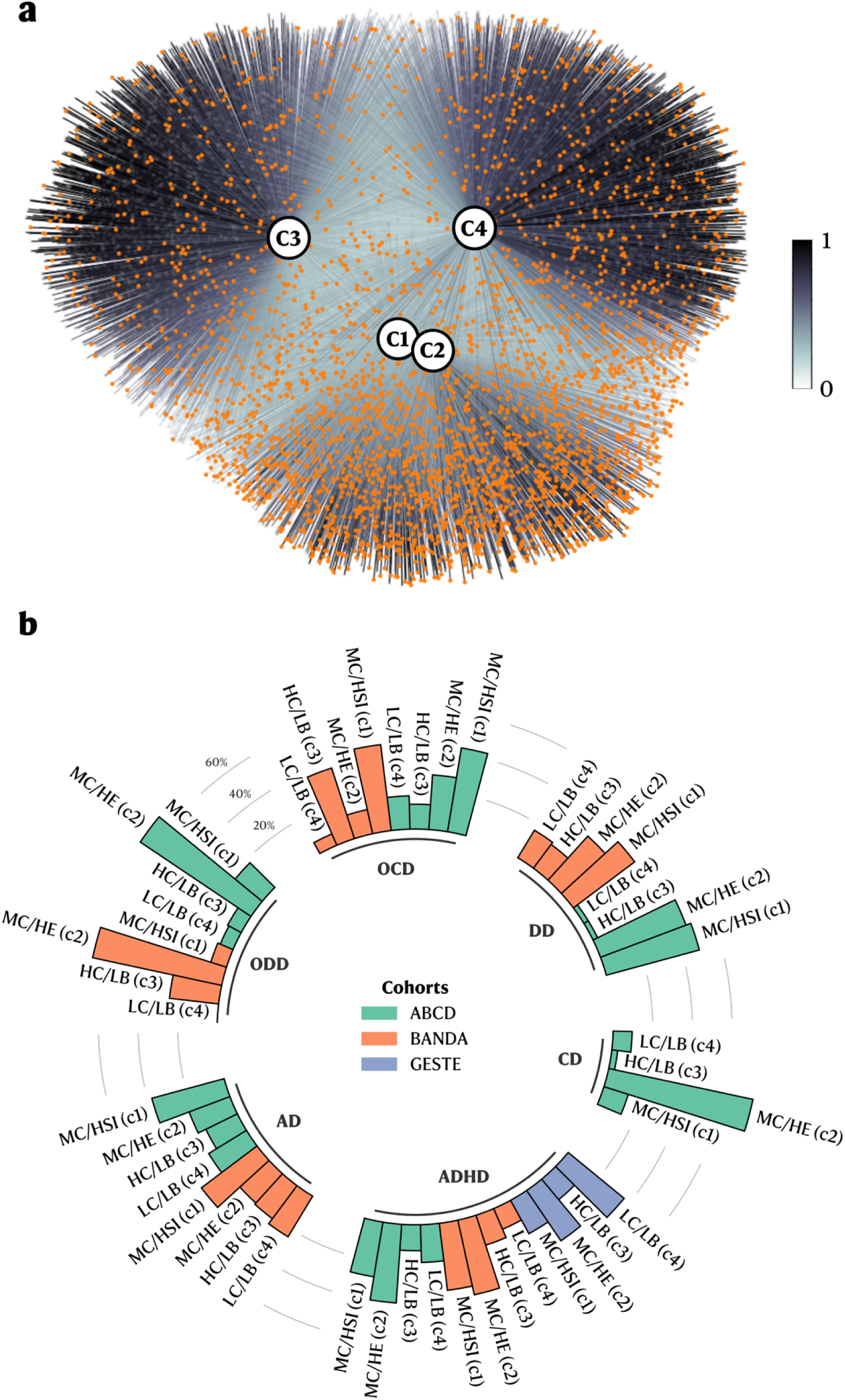
Diagnoses distribution across all profiles. **a.** Graph network including all cohorts (ABCD baseline, BANDA, and GESTE) with participants labeled in orange if they had at least one psychiatric disorder (assessed either via parental K-SADS or from experts). **b.** Circular bar plot showing the percentage of participants with a diagnosis across all the extracted clusters and cohorts. Participants were grouped based on their primary profile (the highest membership value) for the bar plot. AD: Anxiety Disorder. ADHD: Attention Deficit-Hyperactivity Disorder. CD: Conduct Disorder. DD: Depression Disorder. OCD: Obsessive-Compulsive Disorder. ODD: Oppositional Defiant Disorder.

We leveraged graph theory concepts to compute the average weighted shortest path (ASWP) between participants with a specific diagnosis (see Methods). We compared this to a null distribution (e.g., an equally randomly selected number of participants), where significant p-values would reflect a non-random distribution of participants (e.g., they tend to aggregate in specific profiles) (see Methods). Overall, each study and assessment method diagnosis met the FDR-corrected significance threshold for a non-random distribution (Table 2).

**Table 2.** Average shortest weighted path (ASWP) for all diagnoses in the ABCD baseline follow-up, BANDA, and GESTE populations. AD: Anxiety Disorder. ADHD: Attention Deficit-Hyperactivity Disorder. CD: Conduct Disorder. DD: Depression Disorder. OCD: Obsessive-Compulsive Disorder. ODD: Oppositional Defiant Disorder. PSYPATHO: Index indicating participants with at least one psychiatric disorder. *p*_fdr_ : FDR-corrected p-value.

|  | ABCD (parent KSADS) |  | ABCD (youth KSADS) |  | BANDA |  | GESTE |  |
| --- | --- | --- | --- | --- | --- | --- | --- | --- |
| | ASWP | $p_{\text{fdr}}$ | ASWP | $p_{\text{fdr}}$ | ASWP | $p_{\text{fdr}}$ | ASWP | $p_{\text{fdr}}$ |
| **AD** | 0.214 | 0.0002 | 0.217 | 0.0003 | 0.194 | 0.012 | - | - |
| **ADHD** | 0.225 | 0.0002 | - | - | 0.226 | 0.001 | 0.206 | 0.012 |
| **CD** | 0.276 | 0.0002 | - | - | - | - | - | - |
| **DD** | 0.294 | 0.0002 | 0.208 | 0.0004 | 0.212 | 0.001 | - | - |
| **OCD** | 0.234 | 0.0002 | - | - | 0.223 | 0.009 | - | - |
| **ODD** | 0.260 | 0.0002 | - | - | 0.220 | 0.012 | - | - |
| **PSYPATHO** | 0.214 | 0.0002 | 0.213 | 0.0003 | 0.197 | 0.009 | 0.206 | 0.012 |

### Solving challenging real-world questions: impact of environmental factors

The cognitive and behavioral profiles proposed here extract meaningful patterns from the data while retaining all available information from individual participants’ cognitive abilities and behavioral manifestations. To showcase their potential in investigating relationships with external factors, we evaluated the relationship between environmental factors and the cognitive/behavioral profiles in the ABCD and GESTE cohorts. Participants from the BANDA study did not have environmental factors data and were not included in the analysis. One key aspect to consider is the collinearity between each profile membership value. Therefore, we performed two partial least squares regression analyses (PLSR, one for each cohort) to handle collinearity, and we established the significance of both models and coefficients using permutation testing. PLSR returns coefficients that can further be used to understand the influence of environmental factors on the cognitive and behavioral profiles’ membership values (see Methods).

PLSR models were significant in both ABCD (R^2^ = 0.10, *p* < 0.001) and GESTE (R^2^ = 0.06, *p* = 0.002) cohorts. In the ABCD study, participants showing lower school involvement, higher traumatic events, higher prenatal conditions (total conditions and planned pregnancy), higher parental psychopathology, and higher parental education had higher membership value in the MC/HSI profile (Figure 4). Similar trends for school involvement, traumatic events, total prenatal conditions, and parental psychopathology were also predictive of participants in the MC/HE profile, in addition to lower parental acceptance, higher family conflict, and higher school disengagement (Figure 4). Participants’ membership value to profile HC/LB was mainly influenced by higher neighborhood safety, higher parental monitoring, lower family conflict, lower school environment, higher school involvement, lower school disengagement, lower total prenatal conditions, higher birthweight, higher maternal age, lower parental psychopathology, lower sleep hours, and higher parental education (Figure 4). Opposingly, lower parental monitoring, higher school environment, lower school involvement, lower birthweight, and lower parental education were driving higher membership values in profile LC/LB combined with a lower ability to pay medical bills, lower prenatal exposure to drugs, less planned pregnancy, and lower parental psychopathology (Figure 4). In the GESTE study, similar trends were observed. Still, only lower experiences of traumatic events were predictive of participants in profile HC/LB and reached the significance threshold after FDR correction (Figure 4).

**Figure 4.**
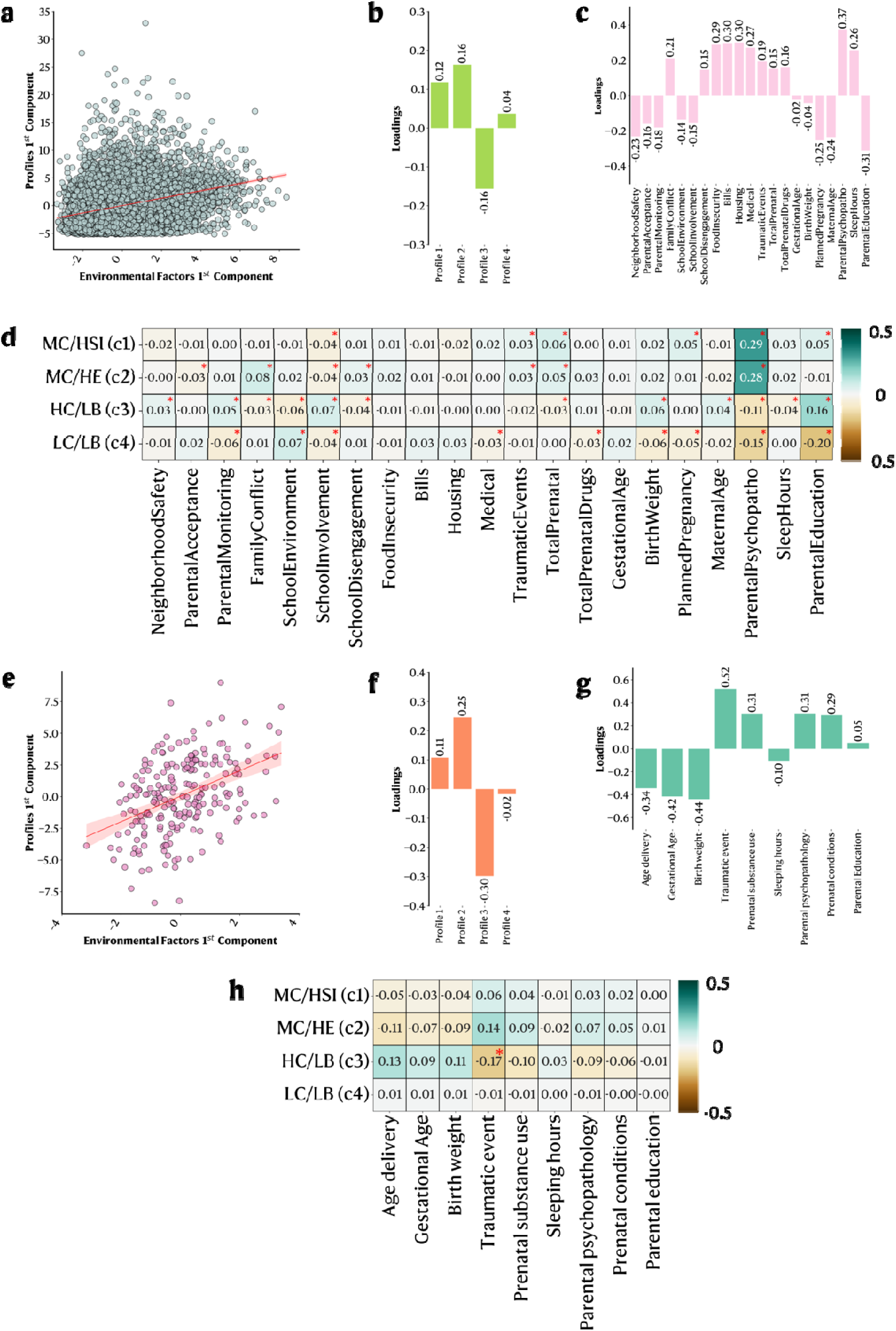
Partial Least Square Regression (PLSR) analysis between environmental factors and cognitive/behavioral profiles in the ABCD and GESTE studies. **a.** Scatter plot of the 1^st^ profile and 1^st^ environmental components in the baseline ABCD cohort. **b.** Loadings of each profile on the 1^st^ component in the ABCD cohort. **c.** Loadings of each environmental factor on the 1^st^ component in the ABCD cohort. **d.** Heatmap of the variables’ coefficients for each profile in the baseline ABCD cohort. **e.** Scatter plot of the GESTE study’s 1st profile and 1st environmental components. **f.** Loadings of each profile on the 1^st^ component in the GESTE cohort. **g.** Loadings of each environmental factor on the 1^st^ component. **h.** Heatmap of the variables’ coefficients for each profile in the GESTE cohort. *: *p*_fdr_ < 0.05.

## Discussion

Although some studies have attempted to extract profiles from populational data, none succeeded in embracing the known continuous nature of cognition and behavior found in the general population. Using data-driven fuzzy clustering, we successfully extracted a 4-profile solution representing the continuum of cognition and behavior in the largest to-date pediatric cohort. Using this referential model, we demonstrated the generalizability of the cognitive and behavioral profiles in two additional independent pediatric cohorts, enabling the direct comparison between studies. Furthermore, we independently reproduced those profiles in subsequent follow-ups within the ABCD study, showcasing their stability across developmental periods.

In all included cohorts and follow-ups, children with moderate cognitive abilities tend to show more behavioral manifestations (externalizing, internalizing, or stress) than children with high or low cognitive abilities. This suggests that the relationship between cognition and behavior might not be linear, significantly contrasting with previously published results^12,13,15,18–20^. However, a recent study questioned that linearity and found that non-linear models were better suited to explain the relationship between cognition and behavior^21^, further supporting our analysis. This contradiction between results highlights the sensitivity of our method since it uncovered the non-linear relationship between cognition and behavior, which other studies could not.

Consistency with existing clinical diagnoses is a key validation step for studies aiming to extract, using data-driven methods, subgroups or profiles from the general population. While it is expected that some differences emerge, considering the known heterogeneity in psychopathology^1,2^, clinical diagnoses remain the gold standard benchmark. Our method was consistent with the existing clinical domains, noting only differences between parent-reported, youth-reported, or clinician-derived diagnoses. Differences between youth-reported and parent-reported diagnoses are known^22^, especially concerning internalizing behavior^23^. Parents often exacerbate their child’s symptoms, whereas youths understate them^23^. However, differences between parent-reported and expert diagnostic opinions are puzzling and suggest substantial implications for research settings, where parent-reported diagnoses are often used as group classifiers^24^. Compared to clinical diagnosis, parental assessment is likely affected by multiple bias sources based on their cultural background and socioeconomic situation. While the current study lacks data and sample size in the expert diagnostic opinions category for a concrete conclusion, it highlights the need for a diagnostic-agnostic approach as proposed here. Our method alleviates this issue by modeling cognition and behavior as non-discrete profiles.

One major current goal in psychiatry is the transition toward precision medicine, which involves including biological and/or environmental factors combined with new data-driven machine-learning methods in the diagnosis process^5,25,26^. To illustrate a study case using our method, we investigated the relationship between our extracted cognitive and behavioral profiles and environmental factors in two of the included cohorts. Our results suggest a strong implication of parental education, lower prenatal substance use, and lower parental psychopathology in cognitive abilities, coherent with previously reported results^11,27^. It further supports the hypothesis that higher-education parents are more likely to invest time (e.g., reading books) and resources in their child’s cognitive development^28^ and might even reinforce the genetic contribution behind cognitive abilities. Unsurprisingly, similar to previous studies, parental psychopathology was highly implicated in profiles showing high behavioral scores (externalizing and internalizing/stress) in addition to prenatal conditions, family conflict, and the experience of traumatic events^11,27^. Indeed, parental behavioral problems are highly predictive of offspring’s psychopathology^29^ and, combined with other environmental factors, are thought to influence the brain’s developmental trajectories^29–33^. Part of this relationship can be explained by the heritability of psychopathology, as shown in genetic studies^34^. Although it is hard to separate the genetic contribution from the environmental factors in the current data, common genetic variants have been shown to have transdiagnostic influences^34^, suggesting fuzzy boundaries even at the genetic level and not only in cognitive/behavioral phenotypes. This demonstrates that our method can derive meaningful relationships between external factors (e.g., environmental factors), cognition, and behavior while retaining all the available information compared to discrete classification methods. Future studies could reuse our method to investigate the underlying neural correlates, as previous studies have reported associations between brain structures and environmental factors^29–33,35,36^.

Our method is ideally suited for a precision medicine framework as it is generalizable and can be used on a single individual. Future research establishing models predicting the trajectories across developmental stages (e.g., are patients moving within those profiles across time? Which profiles show the highest movement across time?) would provide additional information to embody the domain-based framework fully. One crucial advantage of data-driven fuzzy clustering over other methods is the ability to quantify the level of belonging to each profile for each subject. While other methods, such as latent profile analysis, can consider classification uncertainty using probabilities, there are fundamental differences with the proposed approach. Probabilities refer to the likelihood that an event occurs, which, in the case of classification, is the likelihood that a participant is part of a profile. It does not indicate to which degree an individual is part of that profile but quantifies how confident we are with the discrete classification. Fuzzy logic captures the meaning of partial truth, where an individual can simultaneously belong to more than one profile to various degrees (membership values). Those fundamental differences make fuzzy clustering better suited when there is known overlap and when individuals can exhibit characteristics from multiple profiles. It is known that cognition and behavior represent an overlapping continuum of capabilities/manifestations; therefore, embracing the fuzziness in a general population is mandatory and most likely better reflects the real-world scenario compared to subgroup extraction or probabilistic methods.

However, our study also has important limitations to consider. First, not all data collection instruments were identical across studies. While this could limit our reproducibility evaluation in different populations, we mitigated this risk using robust harmonization techniques (see Supplementary Figure 4). Second, the stress variable was unavailable in the GESTE cohort; thus, we imputed it using a sophisticated imputation model and assessed its performance on existing data and against an independent variable closely related to the imputed one (see Supplementary Figures 5 & 6). This imputed variable might have introduced an overestimation of the membership values for profile MC/HSI in the GESTE cohort. Third, we evaluated the relationship with the available environmental factors within our cohorts. However, those factors only capture a subset of the possible environmental exposures. Future studies should include additional variables, such as exposure to substances or diet-related factors, to the ones used in this study. Fourth, all included cohorts were from North American backgrounds (Canada and the USA), which might introduce cultural bias in our results. Future studies should validate the results in other cultural backgrounds.

While this study only examines individual-level relationships, our framework could also be applied to examine population-level relationships, such as the impact of famine and war, which would shift the current profiles’ distribution. This aspect highlights the granularity of the method and the wide range of possible applications. One critical step to attaining precision medicine is the ability to create generalizable frameworks for various populations. Hence, we validated our fuzzy profiles in two smaller independent cohorts (GESTE and BANDA) and across developmental stages using subsequent ABCD follow-ups, demonstrating the generalizability and replicability of the profiles. Additionally, we ensured consistency with existing clinical domains, showing good-to-great concordance and reinforcing the usability of our method in clinical settings. Finally, we showcased how our method could be used to investigate pressing research questions by looking at the relationship between the profiles and environmental factors. Establishing this referential in the ABCD cohort expands the applicability to other smaller cohorts, where statistical power might be insufficient to extract similar profiles.

## Methods

### Study design and participants

ABCD is a multi-site longitudinal prospective cohort of 11,878 children recruited through school systems across 21 sites in the United States^10^. Children aged 9-10 were enrolled in the study during the baseline follow-up from 2016 to 2018. Recruitment strategies were carefully designed to generate a cohort representing the US sociodemographic population distribution. This is a major advantage compared to other studies, as it enhances the ability to identify and study specific neurodevelopmental trajectories. Of the enrolled participants, 10,843 had complete baseline behavioral, cognitive, and psychopathology data and were included in the present primary analysis. Cognitive and behavioral data from the 2-year (n=7,369) and 4-year follow-up (n=2846) were also included to evaluate our extracted profiles’ stability across developmental stages. All data were gathered from the data release 5.1 (more details here: https://wiki.abcdstudy.org/). Further details regarding the participating sites, ethics, study protocols, and investigators are available here: https://abcdstudy.org/.

BANDA is a multi-site prospective cohort of 225 adolescents aged 14-17 recruited through clinics, advertisements (social media, buses, and trains), and newsletters^16^. The study is designed to assess brain differences between three clinical groups: depressed, anxious, and control (defined as participants without any diagnosis). Participants were included in the study if they were fluent in English, between 14-17 years old, eligible for an MRI, and obtained a score higher than 85 on the Wechsler Abbreviated Scale of Intelligence (WASI). Participants were excluded if they had complications at birth, serious medical conditions, history of head injury, prior hospitalization for more than 2 days (neurological or cardiovascular disease), diagnosis of autism spectrum disorder, and used preventive migraine medication daily. Of the enrolled participants, 195 had complete cognitive, behavioral, and psychopathology data from the baseline visit and were included in the present analysis. Further details are available here: https://banda.mit.edu/index.html.

GESTE is a population-based cohort in Sherbrooke, Quebec, Canada^17^. Data used in the present study comes from the fourth follow-up in which children aged 9-13 underwent complete neuropsychological assessment (n = 309 participants). Initial enrollment (n = 800) happened between 2007 and 2009 during the first trimester or at birth if the participant’s mother met the following criteria: 1) healthy women over 18 years old without severe preterm birth and 2) no chronic medical conditions. Of the participants seen during the fourth follow-up, 271 children with available behavioral, cognitive, and psychopathology data were included in the present analysis. All study protocols were approved by both the Institutional Ethics Boards of the University of Sherbrooke and Columbia University. All population demographics are presented in Table 1.

### Procedures

During the baseline follow-up, ABCD participants underwent an exhaustive neurocognitive battery (further described in Luciana et al., 2018) comprising the NIH Toolbox cognition measures (NIHTB)^38^, the Little Man’s Task (LMT)^39^, the Rey Auditory Verbal Learning Test (RAVLT)^37^, and the Wechsler Intelligence Test for Children-V (WISC-V) Matrix Reasoning task^40^. ABCD participants were readministered the NIHTB^38^ and the LMT^39^ combined with the RAVLT^37^ and the Game of Dice (DICE) task^41^ during the 2-year follow-up and the DICE^41^ and the Behavioral Indicator of Resiliency to Distress (BIRD) task^42^ during the 4-year follow-up. BANDA participants underwent a similar neurocognitive battery comprising the NIH Toolbox^38^, the University of Pennsylvania Computerized Neuropsychological Test Battery (Penn Test Battery)^43^, and the Weschler Abbreviated Scale of Intelligence 2^nd^ edition (WASI-II)^44^. For both ABCD and BANDA participants, each test was administered using a computerized version. During the fourth follow-up, at an in-person visit, GESTE participants completed 7 subtests of the Wechsler Intelligence Test for Children-V (WISC-V) test battery^40^. For each study, uncorrected scaled scores were used in further analyses. Following Moore & Conway, 2023, split-sample sequential exploratory factor analysis (EFA) and confirmatory factor analysis (CFA) were performed to obtain latent factors representing three major cognitive domains in each study: verbal ability (VA), executive functions/processing speed (EF/PS), and memory (MEM). Further details regarding the administration, loadings, and fit indices for the EFA/CFA models are available in Supplementary Material S6, S7, & S8, and Tables 2, 3, 4, 5, 6, 7, 8, 9, 10, and 11. The mean scaled values of the three latent factors are presented in Table 1.

The parents or caregivers of the ABCD and BANDA participants answered the Child Behavioral Checklist (CBCL) based on six months before the follow-up^45^. Using the syndrome and 2007 CBCL scales, scores for internalizing (sum of the anxious/depressed, withdrawn/depressed, and somatic complaints variables), externalizing (sum of the rule-breaking and aggressive behavior variables), and stress problems were computed and included as behavioral indicators. Parents or caregivers of the GESTE participants were asked to complete the Behavior Assessment System for Children 3^rd^ edition (BASC3) parent rating scales. The BASC3 questionnaire is designed to inform on the child’s behavior across four composite scores: externalizing problems (sum of hyperactivity, aggression, and conduct problems subscales), internalizing problems (sum of anxiety, depression, and somatization subscales), behavioral symptoms index (sum of atypicality, withdrawal and attention problems subscales), and adaptive skills (sum of adaptability, social skills, leadership, activities of daily living, and functional communication subscales)^46^. We retained only the externalizing and internalizing composite scores to ease the comparison between studies. Throughout this article, we refer to those variables (externalizing, internalizing, and stress) as “behavioral scores”.

In addition to cognitive and behavioral data, we included available environmental factors encompassing perinatal conditions (maternal age, gestational age, birth weight, substance use, maternal medical conditions, and planned pregnancy), adverse childhood experiences (ACEs; traumatic events, family conflict, and parental psychopathology), sleep hours, neighborhood safety, school factors (environment, disengagement, and involvement), parental factors (acceptance, monitoring, and education level), and economic factors (ability to pay bills, provide food, housing, and medical care) from the ABCD study^47^. Available variables from the GESTE study included the perinatal conditions, history of traumatic events, sleep hours, and history of parental psychopathology. Due to the unavailability of such variables in the BANDA study, those participants were excluded from this analysis. Detailed descriptions of variables, instruments, and encoding methods are presented in Supplementary Material S2 and Supplementary Table 1.

A computerized version of the Kiddie Schedule for Affective Disorders and Schizophrenia for School-Aged Children (KSADS) completed by the parent or caregiver was used to assess multiple aspects of the youth’s psychopathology within the ABCD and BANDA study. The KSADS is a widely used tool that highly correlates with DSM-V criteria and has been proven helpful in clinical settings and research studies^24^. Using a similar approach to previous work by Bernanke et al., 2022, we extracted and created categorical diagnosis variables for anxiety disorder (AD), attention deficit-hyperactivity disorder (ADHD), conduct disorder (CD), depressive disorder (DD), obsessive-compulsive disorder (OCD), and oppositional defiant disorder (ODD) (see Supplementary Material S3 for more details). In addition to the parent-administered KSADS, we extracted diagnosis variables from the youth-administered KSADS version from ABCD baseline follow-up. Due to the limited number of modules administered, we only extracted AD and DD categorical diagnosis variables (see Supplementary Material S3 for more details). Diagnosis information from the GESTE participants was pulled directly from the medical records. This article will use the term “psychopathology” to refer to those diagnoses derived from both medical records and questionnaires. Diagnosis distributions are presented in Table 1.

### Statistical analyses

The Supplementary Material S9-10 provides a complete description of the preprocessing and statistical analyses, while an overview is presented in Figure 1. Briefly, data from each cohort and follow-up were harmonized (Supplementary Figures 1, 2, 3, and 4) and residualized for covariates (age, sex, ethnicity, and handedness). We imputed the missing Stress variable in the GESTE cohort using a K-Nearest Neighbor model. The ABCD cohort was used as the discovery sample for all analyses, and both remaining cohorts were used to validate and replicate the results. We applied the fuzzy C-Means clustering algorithm to create cognitive and behavioral profiles within the ABCD study as a reference dataset. The optimal number of clusters was derived using the silhouette score (Supplementary Material S10 and Supplementary Figure 7). To enable the comparison between studies, we used the ABCD clusters’ centroids to predict the membership values for each BANDA and GESTE participant. Prediction using the centroids does not require additional studies (used in prediction) to cover all the original extracted profiles or to have a similar sample size. Briefly, using the cognitive and behavioral scores of the “new” participants, the prediction model will “map” the participants by computing the distance from each centroid and then returning the membership values (more details in Supplementary Material S10). A one-way ANOVA followed by a Tukey Honestly Significant Difference (HSD) post hoc test was performed to evaluate the difference in means between profiles and cohorts. When comparing scores or values between profiles, we used the participants’ primary cluster (the highest membership value) as the categorization criteria. Additional FCM analyses were independently performed (not predicted) for each ABCD follow-up to evaluate the consistency of the extracted profiles across developmental stages using the same preprocessing steps (harmonization and residualization). The extracted profiles were assessed using the same methods as the baseline data. Clusters were visualized by constructing a weighted graph network using the participants as nodes and membership values as the edge’s weight^48^. To quantify the non-random distribution of psychopathology on the graph network, we computed the average shortest weighted path (ASWP) between all nodes of interest. A higher value translates to a more compact aggregation and a non-random distribution. Permutation testing using 5,000 iterations was performed to estimate the results’ significance. We conducted two partial least square regression analyses (PLSR) (one for each study) using environmental factors as predictors and the profiles’ membership values as dependent variables. Membership values for each profile were included as continuous variables, leveraging the ability of PLSR to handle multicollinearity. PLSR extracts components for predictors and dependent variables to maximize the covariance between those two sets and returns coefficients for each predictor to each dependent variable. Those coefficients can be used to understand the influence of the predictors on the dependent variables. For example, while it does not reflect a direct linear relationship due to the projection into latent space in PLSR, a high coefficient for a predictor will reflect a higher dependent variable if that predictor’s value increases. Ten-fold cross-validation was used to assess the optimal number of components. Permutation testing using 10,000 iterations assessed the model and coefficients’ significance. All reported p-values are corrected for false discovery rate (FDR), and the post-correction significance threshold was set to *p*_fdr_ < 0.05^49^. Complete code to reproduce the analyses is available here: https://github.com/Labo-MAB/Gagnon_FuzzyProfiles_2025. A Python package containing general-use command-line scripts allowing researchers to use this method in new populations is available here: https://github.com/gagnonanthony/NeuroStatX.

## Data Availability

Adolescent Brain Cognitive Development (ABCD) and Boston Adolescent Neuroimaging of Anxiety and Depression (BANDA) data are available online at https://nda.nih.gov/ after obtaining a data use certificate. Data for the GESTation and Environment (GESTE) study are available upon reasonable request to the authors.

## Contributors

AG, MAB, MD, and LT contributed to the study design and conceptualization. LT, JL, AB, and JP acquired funding for the GESTE cohort. AG, VG, and AD were involved in data collection and curation and had direct access to the data. AG wrote the final draft. All authors contributed to the interpretation of findings and approved the final version of the manuscript before submission. AG and LT were responsible for the decision to publish.

## Data Sharing

The Adolescent Brain and Cognitive Development (ABCD) and the Boston Adolescent Neuroimaging of Depression and Anxiety (BANDA) are freely available datasets accessible via the NIMH Data Archive platform (https://nda.nih.gov/). Researchers must apply for a Data Use Certificate, which will be reviewed and granted by study administrators. Data for the GESTation and Environment cohort is not publicly available but can be accessed by contacting the principal investigator in charge of the study, Dr. Larissa Takser. Code to reproduce the findings, including data curation, preprocessing, analysis, and visualization, is accessible at https://github.com/Labo-MAB/Gagnon_FuzzyProfiles_2025.

## Acknowledgments.

Data used in the preparation of this article were obtained from the Adolescent Brain Cognitive Development^SM^ (ABCD) Study (https://abcdstudy.org), held in the NIMH Data Archive (NDA). This is a multisite, longitudinal study designed to recruit more than 10,000 children age 9-10 and follow them over 10 years into early adulthood. The ABCD Study® is supported by the National Institutes of Health and additional federal partners under award numbers U01DA041048, U01DA050989, U01DA051016, U01DA041022, U01DA051018, U01DA051037, U01DA050987, U01DA041174, U01DA041106, U01DA041117, U01DA041028, U01DA041134, U01DA050988, U01DA051039, U01DA041156, U01DA041025, U01DA041120, U01DA051038, U01DA041148, U01DA041093, U01DA041089, U24DA041123, U24DA041147. A full list of supporters is available at https://abcdstudy.org/federal-partners.html. A listing of participating sites and a complete listing of the study investigators can be found at https://abcdstudy.org/consortium_members/. ABCD consortium investigators designed and implemented the study and/or provided data but did not necessarily participate in the analysis or writing of this report. This manuscript reflects the views of the authors and may not reflect the opinions or views of the NIH or ABCD consortium investigators. The ABCD data repository grows and changes over time. The ABCD data used in this report came from <u>10.15154/z563-zd24</u>. DOIs can be found at https://nda.nih.gov/abcd/abcd-annual-releases. Data were provided by the Boston Adolescent Neuroimaging of Anxiety and Depression (BANDA) Consortium’s Human Connectome Project, supported by 1U01MH108168 (PIs: Susan Whitfield-Gabrieli, John Gabrieli). Data was also provided by the GESTE study, supported by the National Institute of Environmental Health (grant # R01ES027845). The authors thank all GESTE participants and staff members for their dedication and hard work. AG was supported by a Canadian Institute of Health Research Doctoral Award (#493956). MAB is supported by a Junior 1 career award from the Fonds de Recherche du Québec – Santé (FRQS).

