## Supplement for "Beyond Discrete Classifications: A Computational Approach to the Continuum of Cognition and Behavior in Children"

**Gagnon A, et al.**

**Table of Contents**

| S1 | Neurocognitive measures and administration. | page 4 |
| --- | --- | --- |
| S2 | Extraction of the environmental factors from the ABCD cohort. | page 5 |
| S3 | Extraction and creation of the categorical diagnosis variables from the Kiddie Schedule for Affective Disorders and Schizophrenia for School-Aged Children (KSADS). | page 5 |
| S4 | Data harmonization across ABCD sites. | page 7 |
| S5 | Residualization for covariates in all studies. | page 7 |
| S6 | Split-sample EFA and CFA to extract the latent structure of cognition within the ABCD study. | page 8 |
| S7 | Split-sample EFA and CFA to extract the latent structure of cognition within the BANDA study. | page 9 |
| S8 | Split-sample EFA and CFA to extract the latent structure of cognition within the GESTE study. | page 10 |
| S9 | Data harmonization and imputation of the stress problems score within the GESTE study. | page 11 |
| S10 | Fuzzy clustering method. | page 12 |
| S11 | Predicting the membership values for BANDA and GESTE using the ABCD cluster’s centroids. | page 13 |

**Supplementary Tables**

| Table S1 | List of all measures and coding methods to extract environmental factors from the ABCD study. | page 17 |
| --- | --- | --- |
| Table S2 | Latent factor loadings for all variables and associated eigenvalues for the baseline ABCD study. | page 20 |
| Table S3 | ABCD baseline split-sample Confirmatory Factor Analysis (CFA) coefficients. | page 20 |
| Table S4 | Latent factor loadings for all variables and associated eigenvalues for the 2-year follow-up ABCD study. | page 21 |
| Table S5 | ABCD 2-year follow-up split-sample Confirmatory Factor Analysis (CFA) coefficients. | page 21 |
| Table S6 | Latent factor loadings for all variables and associated eigenvalues for the 4-year follow-up ABCD study. | page 22 |
| Table S7 | ABCD 4-year follow-up split-sample Confirmatory Factor Analysis (CFA) coefficients. | page 22 |
| Table S8 | Latent factor loadings for all variables and associated eigenvalues for the BANDA study. | page 23 |
| Table S9 | BANDA split-sample Confirmatory Factor Analysis (CFA) coefficients. | page 23 |
| Table S10 | Latent factor loadings for all variables and associated eigenvalues for the GESTE study. | page 24 |
| Table S11 | GESTE split-sample Confirmatory Factor Analysis (CFA) coefficients. | page 24 |
| Table S12 | Demographic information for the 2-year and 4-year ABCD samples. | page 25 |
| Table S13 | One-way ANOVA and Tukey HSD post hoc test results for each cognitive and behavioral symptoms variable in the ABCD baseline. | page 26 |
| Table S14 | One-way ANOVA and Tukey HSD post hoc test results for each cognitive and behavioral symptoms variable in the BANDA study. | page 26 |
| Table S15 | One-way ANOVA and Tukey HSD post hoc test results for each cognitive and behavioral symptoms variable in the GESTE study. | page 26 |
| Table S16 | One-way ANOVA and Tukey HSD post hoc test results for each cognitive and behavioral symptoms variable in the ABCD 2-year follow-up. | page 27 |
| Table S17 | One-way ANOVA and Tukey HSD post hoc test results for each cognitive and behavioral symptoms variable in the ABCD 4-year follow-up. | page 27 |
| Table S18 | One-way ANOVA and Tukey HSD post hoc test results between each cohort mean scores and each profile. | page 28 |

**Supplementary Figures**

| Figure S1 | Boxplot of the ABCD baseline site’s distributions for each cognitive and behavioral variable before and after harmonization. | page 29 |
| --- | --- | --- |
| Figure S2 | Boxplot of the ABCD 2-year follow-up site’s distributions for each cognitive and behavioral variable before and after harmonization. | page 30 |
| Figure S3 | Boxplot of the ABCD 4-year follow-up site’s distributions for each cognitive and behavioral variable before and after harmonization. | page 31 |
| Figure S4 | Boxplot of the common variables distribution between the ABCD, GESTE, and BANDA study before and after harmonization using the ComBat framework. | page 32 |
| Figure S5 | Evaluation of the performance of the k-Nearest Neighbors imputation (KNN) model using the Euclidean distance as weight. | page 33 |
| Figure S6 | Scatter plot of the imputed stress problems score compared to the BASC-3 Depression composite score reflecting stress and general unhappiness and sadness. | page 34 |
| Figure S7 | Line plot of the silhouette score for each number of clusters for each ABCD follow-up (baseline, 2-year, and 4-year). | page 35 |
| Figure S8 | FCM clustering results with radar plot for each study. | page 36 |
| Figure S9 | Diagnosis distribution across all profiles for each independent cohort. | page 37 |
| Figure S10 | Youth KSADS diagnosis distribution across all profiles for the ABCD baseline data. | page 38 |

**Data Collection**

1. *Neurocognitive measures and administration.*

As described in Luciana et al., 2018, ABCD participants underwent a neurocognitive battery comprising five tests: NIH Toolbox (NIHTB), Little Man’s Task (LMT), Rey Auditory Verbal Learning Test (RAVLT), and Wechsler Intelligence Test for Children-V (WISC-V) Matrix Reasoning task. The NIHTB comprises six tests designed to evaluate episodic memory, executive function, attention, working memory, processing speed, and language abilities and was administered using an iPad ^1,2^. The LMT is a computerized test measuring the visuospatial reasoning ability, in which participants must determine which hand a man is holding his briefcase ^1,3^. Amongst the test scores, the percentage of total correct answers was retained and used throughout this manuscript. The RAVLT contains seven trials assessing auditory learning, short- and long-term memory, and recognition. It was administered using a computerized version on an iPad using the Q-Interactive Pearson Assessment platform ^1,4^. Like other articles, the total correct score for the longer-term memory trial was used in the analyses ^5,6^. A computerized version of the WISC-V matrix reasoning subtest was administered to assess non-verbal reasoning, a high predictor of fluid reasoning and general intellectual ability ^1,7^. Introduced for the 2-year follow-up, ABCD participants were administered the Game of Dice task ^8^. The DICE is designed to measure decision-making abilities under various risk conditions. Participants are asked to predict the result of a dice roll from four possibilities ranked from more risky to less risky based on their probabilities of happening. Payoffs are associated with each option according to the probability of one to happen (lower probability = higher reward). Results are collected from over 18 trials to assess the risk-taking of each participant. Introduced in the 4-year follow-up, participants underwent the Behavioral Indicator of Resiliency to Distress (BIRD) task. The task aims to assess resilience under distress by emitting an unpleasant sound when the participant fails to reach the objective (reaching a green dot) in time. At every point, the participant is allowed to quit the task. Uncorrected scaled scores were used when applicable. The neurocognitive assessment in the BANDA study comprises subtests from the NIH Toolbox, the University of Pennsylvania Computerized Neuropsychological Test Battery (Penn Test Battery), and the Weschler Abbreviated Scale of Intelligence 2^nd^ edition (WASI-II) ^2,9,10^. All subtests were administered using a computerized version for a total duration of 75 minutes. The uncorrected scaled scores were extracted and used in this analysis when appropriate. As part of the fourth follow-up, GESTE participants used pen and paper to complete the Wechsler Intelligence Test for Children-V (WISC-V) test battery. It consisted of seven subtests assessing verbal reasoning, verbal comprehension, perceptual reasoning, working memory, and processing speed ^11^. The uncorrected scaled scores were extracted and used in the analyses.

2. *Extraction of the environmental factors from the ABCD cohort.*

To create the environmental factors used in the partial least square regression (PLSR) analysis, we extracted variables from several data instruments in ABCD Release 5.1. All variable details and original encoding can be seen here: <https://data-dict.abcdstudy.org/>. The Supplementary Table 1 provides the encoding details for each environmental factor. For GESTE participants, perinatal factors were pulled directly from medical records, while adverse childhood experiences (ACEs) and parental education were part of the sociodemographic questionnaire.

3. *Extraction and creation of the categorical diagnosis variables from the Kiddie Schedule for Affective Disorders and Schizophrenia for School-Aged Children (KSADS).*

The KSADS is a widely used tool that highly correlates with current DSM-V criteria and has been proven useful in multiple clinical and research settings ^12^. It reports on multiple diagnoses regarding the current and past manifestation of symptoms. Administration is structured in three phases: 1) an unstructured introductory interview, 2) a diagnosis screening interview, and 3) additional modules complementing the diagnosis criteria. Multiple variables regarding past presentation, present manifestation, and partial remission of a diagnosis are reported in the KSADS report. Within the ABCD study, following the work Bernanke et al., 2022, we combined unspecified Attention Deficit-Hyperactivity Disorder (ADHD), partial remission ADHD, present ADHD, and past ADHD to create a single ADHD diagnosis variable. Similarly, we combined panic disorder, agoraphobia, separation anxiety, social anxiety, and generalized anxiety disorder variables to create the anxiety disorder (AD) diagnosis variable. The depressive disorder diagnosis variable (DD) represents the combination of present and partial remission persistent depressive disorder and present and partial remission major depressive disorder. The single variables representing the present manifestations of obsessive-compulsive disorder (OCD), oppositional defiant disorder (ODD), and conduct disorder (CD) were used for those specific diagnoses. To compare with the parent-administered KSADS, we also extracted diagnosis variables from the youth-administered KSADS. Only the KSADS modules assessing AD and DD were administered during the baseline follow-up. Using a similar approach to the parent-administered KSADS, we combined social anxiety and generalized anxiety disorder (only ones available) into a single youth-reported AD categorical variable. For the DD categorical variable, we combined persistent depressive disorder past/present and present/partial remission major depressive disorder.

The KSADS was also administered within the BANDA study using a previous version that did not include DSM-V diagnosis criteria. The study investigators reviewed all modules to validate the consistency with DSM-V criteria and provide a DSM-V-adapted KSADS ^13^. Thus, we combined past/current ADHD, current/lifetime other specified ADHD, and current/lifetime unspecified ADHD variables to create a single ADHD diagnosis variable. For the AD variable, we combined past/current generalized anxiety disorder, agoraphobia, separation anxiety, panic disorder, social phobia, illness anxiety, other specified anxiety, and unspecified anxiety. The DD diagnosis variable comprises past/current major depressive disorder and other specified depression disorders; meanwhile, the OCD diagnosis variable combines past/current OCD, other specified OCD, and unspecified OCD. The ODD diagnosis variable combines ODD's current/past manifestations. The CD diagnosis variable comprises past/current CD, other specified, unspecified disruptive impulse-control, and CD variables. For both studies, the final diagnosis variables included ADHD, AD, DD, OCD, ODD, and CD.

**Statistical Analysis and Data Preprocessing.**

4. *Data harmonization across ABCD sites.*

Since ABCD is a multi-site study spanning across the United States, it is essential to consider variations in the administration of data collection instruments. Those small variations introduce non-biological effects in the data that might influence the results of a specific analysis. To control for those effects, we harmonized the cognitive and behavioral data across the ABCD sites using the ComBat framework ^14,15^. Specifically, we used the neuroCombat Python package ^14^ with the empirical Bayes method combined with parametric adjustments. Harmonization was carried out before any processing steps. Visual representations of the site’s distributions before and after harmonization for each cognitive and behavioral variable and each follow-up are presented in Supplementary Figures 1, 2, & 3.

5. *Residualization for covariates in all studies.*

To control for potential confounders, we residualized, after harmonization across sites, the raw behavioral and cognitive scores by fitting a linear regression model using age, sex, ethnicity, and handedness as predictors and each score as the dependent variable (var ~ age + sex + ethnicity + handedness). This step was performed independently in all studies, for all follow-ups (for ABCD), and the residualized values were kept for following processing steps and analysis. Thus, by removing the effect of age, sex, ethnicity, and handedness, we can perform a clustering analysis that will parse the data according to true cognitive and behavioral patterns.

6. *Split-sample EFA and CFA to extract the latent structure of cognition within the ABCD study.*

Following previous work Moore & Conway, 2023 and Thompson et al., 2019, we extracted latent factors that summarize the structure of cognitive abilities using a split-sample EFA and CFA approach. For the baseline follow-up, raw variables included Matrix Reasoning, Little Man’s Task total percentage of correct answers (LMT), long-term memory total correct score from the Rey Auditory Verbal Learning Task (RAVLT), Oral Reading Recognition (ORR), Pattern Sequence Memory Task (PSMT), Pattern Comparison Processing Speed (PCPS), Dimensional Change Card Sorting (DCCS), List Sorting Working Memory (LSWM), Flanker Inhibitory Control & Attention (FICA), and Picture Vocabulary Task (PVT). Uncorrected scaled scores were used when available. The total sample (n = 10,843) was randomly split into two independent samples. EFA with a 3 latent factor (determined by parallel analysis) solution was performed on the first independent sample using an oblimin rotation and minimum residual solution. The returned factor structure was fairly similar to previous studies: 1) verbal ability (VA) comprising mostly PVT, ORR, and LSWM, 2) Executive function and processing speed (EF/PS) comprising FICA, DCCS, and PCPS, and 3) memory (MEM) comprising PSMT, LSWM, and RAVLT ^5,6^. Loadings and explained variance are presented in Supplementary Table 2. Retaining only those variables for each latent factor, a CFA was performed using the χ^2^, the comparative fit index (CFI), the Tucker-Lewis Index (TLI), and root mean squared error approximation (RMSEA) as goodness-of-fit indicators. A significant χ^2^, a CFI and TLI > 0.95, and an RMSEA < 0.05 are excellent fit indices for a CFA model ^16–18^. The fitted model returned a χ^2^ of 136.46 (*p* < 0.001), a CFI of 0.986, a TLI of 0.976, and an RMSEA of 0.037. The model’s coefficients, standard errors, Z-values, and p-values are presented in Supplementary Table 3. For the 2-year follow-up, included raw variables were the LMT percentage of correct answers, RAVLT long-term memory total correct score, FICA, PVT, PCPS, PSMT, ORR, and DICE net score (defined as the number of safe bets minus the number of risky bets). The same split-sample method was applied to the 2-year follow-up. The returned factor structure was similar to the baseline follow-up: 1) VA: PVT and ORR, 2) EF/PS: FICA and PCPS, and 3) MEM: PSMT and RAVLT (Supplementary Table 4). Fitting those variables within a CFA model returned excellent fit indicators (χ^2^ = 49.27 (*p* < 0.001), CFI = 0.987, TLI = 0.969, and RMSEA = 0.044). Coefficients, standard errors, Z-values, and p-values are presented in Supplementary Table 5. For the 4-year follow-up, the variables included were the LMT percentage of correct answers, FICA, PVT, PCPS, LSWM, PSMT, ORR, DICE net score, and BIRD score. As previously, EFA analysis returned a three-factor solution: 1) VA: PVT and ORR, 2) EF/PS: FICA and PCPS, and 3) MEM: LSWM and PSMT (Supplementary Table 6). As for the other follow-ups, the CFA model returned excellent fit indices (χ^2^ = 22.28 (*p* = 0.001), CFI = 0.990, TLI = 0.976, and RMSEA = 0.044). Details are presented in Supplementary Table 7. All analyses were performed in Python using the NeuroStatX toolbox (<https://github.com/gagnonanthony/NeuroStatX.git>) and the Semopy package ^19^.

7. *Split-sample EFA and CFA to extract the latent structure of cognition within the BANDA study.*

We performed a split-sample EFA and CFA to extract the latent cognitive structure of the BANDA study. Raw variables included five subtests from the NIH toolbox (ORR, LSWM, DCCS, FICA, and PCPS) ^2^, two subtests from the Penn Test Battery (Working Memory (PennWM), and Matrix Reasoning (PennMR)) ^9^, and the WASI-II Vocabulary subtest (WASI-VC) ^10^. The total sample with available complete cognitive data (n = 195 participants) was randomly separated into two independent training and test samples. EFA was initially performed using the oblimin rotation and the minimal residual solution to extract a three-latent factor structure determined by parallel analysis. The obtained factor structure was as follows: 1) VA: ORR, WASI-VC, and LSWM, 2) EF/PS: DCCS, FICA, and PCPS, and 3) MEM: PennMR, PennWM, and LSWM. Loadings, communalities, and eigenvalues are presented in Supplementary Table 8. Retaining only those variables for each latent factor, we performed a CFA using the χ^2^, CFI, TLI, and RMSEA as fit indicators. The model returned a χ^2^ = 27.31 (*p* = 0.038), a CFI of 0.919, a TLI of 0.859, and an RMSEA of 0.085. While those results don’t indicate a perfect fit to the sample but rather a good fit, this structure was kept to facilitate the comparison between each study included in the present analysis. CFA coefficients, standard errors, Z-values, and p-values are presented in Supplementary Table 9.

8. Split-sample EFA and CFA to extract the latent structure of cognition within the GESTE study.

We conducted a split-sample sequential EFA and CFA to extract the latent cognitive structure of the GESTE study. Raw variables included in the analysis were: WISC-V Block (BL), WISC-V Similarities (SI), WISC-V matrix reasoning (MR), WISC-V digit span (DS), WISC-V code (CO), WISC-V verbal comprehension (VC), and WISC-V balance (BA). Uncorrected scaled scores were used when available. The total sample (n = 271 participants) was randomly separated into two independent samples. EFA was performed to extract initial loading values using a three-latent factor structure (determined by parallel analysis) using the same rotation (oblimin) and solution as for the ABCD study. The final returned factor structure was defined as 1) VA: SI, and VC, 2) MEM: BL, MR, DS, and BA, and 3) EF/PS: CO. Returned loadings and eigenvalues are presented in Supplementary Table 10. Retaining only those variables for each factor, we conducted a CFA using the χ^2^, CFI, TLI, and RMSEA as fit indicators. The model returned a χ^2^ = 11.66 (*p* = 0.390), a CFI of 0.997, a TLI of 0.993, and an RMSEA of 0.021, indicating a great fit with the underlying data. This structure was kept to be consistent with the other cohorts. CFA coefficients, standard errors, Z-values, and p-values are presented in Supplementary Table 11.

9. *Data harmonization and imputation of the stress problems score within the GESTE study.*

Using the ComBat framework ^14,15^, we harmonized the internalization, externalization, VA, EF/PS, and MEM variables distributions using the ABCD study as the reference. The harmonization computation was performed using the neuroCombat Python package ^14^ with the empirical Bayes method combined with parametric adjustments. The variable’s distributions before and after harmonization are presented in Supplementary Figure 4. Following harmonization, we trained a k-Nearest Neighbors (KNN) imputation model using the six variables within the ABCD study (internalization, externalization, stress, VA, EF/PS, and MEM). We independently ran multiple imputations of known variables in the GESTE study to determine the optimal number of neighbors to include in the prediction (Supplementary Figure 5.). We then evaluated the correspondence between the imputed and real scores using Pearson’s correlation coefficient. From the tested number of neighbors (5, 10, 20, 30, 40, 50, 60, 70, 80, 90, 100, 150, 200, 250, 300, 350, and 400), the use of 100 neighbors to impute the scores returned the highest Pearson’s correlation coefficient within the GESTE study while minimizing the number of neighbors used (Supplementary Figure 5.). Thus, the final imputation of the stress problems score in the GESTE study was performed using 100 neighbors weighted by the Euclidean distance. To confirm the validity of the imputed stress problems score in the GESTE cohort, we evaluated the correlation between the imputed values and the depression composite score from the Behavioral Assessment System for Children 3^rd^ edition (BASC-3) parent rating scales, which reflect stress problems combined with general feelings of unhappiness and sadness ^20^. As expected, Pearson’s correlation coefficient showed a high correlation (*r* = 0.83, *p* < 0.001) between the imputed stress problems score and the BASC-3 depression score (Supplementary Figure 6.).

10. *Fuzzy clustering method.*

The main advantage of using fuzzy clustering over classical clustering algorithms is the ability to extract membership values for each subject to each cluster. Within this framework, each subject belongs partially to multiple profiles simultaneously, allowing for the natural continuous distribution of cognition and behavior. Quantifying the membership to a profile represents more real-world scenarios than discrete classification within categorical groups. Therefore, we applied a fuzzy C-Means (FCM) algorithm using the Mahalanobis distance metric to create cognitive and behavioral profiles within the ABCD study as a reference dataset. The optimal number of clusters was derived using the silhouette score. The silhouette score helps quantify the consistency within clusters, ranging from -1 to 1, where high positive values denote a good similarity within his cluster and good separability from data within other clusters. We used each subject's main profiles (profiles with the highest membership value) to derive a mean silhouette score for all the samples (Supplementary Figure 7). Following FCM, each subject is assigned a new set of features (*M_1_*, …, *M_k_*) for *k* clusters representing the membership values to each extracted profile. Following FCM on the ABCD dataset, the centroids of all clusters were used to predict membership values for both the GESTE and BANDA studies. Using the centroids from the ABCD fuzzy clustering, the prediction model repeats the clustering process without updating the centroids’ location. This allows us to find the membership values within the latent space at all points. Since this is a deterministic process and does not update based on the predicted new data, additional cohorts do not need a high sample size or complete coverage of all the initially extracted profiles in the discovery dataset. The distance from each centroid is computed and used to assess the membership values for the new participant. Using the highest membership value as the primary cluster, we performed a one-way ANOVA to establish significant differences in means for each cognitive and behavioral score between the extracted profiles. Data from each ABCD follow-up were independently processed by repeating the same FCM analysis and subsequent evaluation methods, including the harmonization and residualization preprocessing steps (Supplementary Figures 2 and 3). We visualized the profiles by building an undirected, weighted graph network with the subjects as nodes and membership values as the edge’s weight ^21^. The graph layout was then computed using the Frutcherman-Reingold force-directed algorithm. To quantify the non-random distribution of subjects within the network, we used the average shortest weighted path metric (ASWP) between all nodes of interest using the membership values as weight. A higher value translates to a more compact aggregation and a non-random distribution. To ensure the significance of the results, we computed a null distribution using 5,000 permutations of randomly selected nodes for each hypothesis. We compared the value to this distribution using a single-tailed parametric t-test. The resulting p-values were FDR-corrected ^22^.

**Supplementary Results.**

11. *Predicting the membership values for BANDA and GESTE using the ABCD cluster’s centroids.*

Since performing independent FCM analysis within both remaining cohorts is impossible due to the limited sample size in each study, we predicted the membership values for BANDA and GESTE using the ABCD cluster’s centroids. Combining the BANDA, GESTE, and ABCD membership values, we overlaid the BANDA and GESTE participants on the ABCD graph network file to evaluate the network coverage for both cohorts. Participants were distributed evenly across the graph network, suggesting a good coverage of all profiles in both cohorts (Supplementary Figure 8A). Computing the mean values and standard deviations stratified by clusters for all cognitive and behavioral variables returned the same pattern across datasets. Profiles C3, C4, and C7 represent low behavioral scores with high (HC/LB), low (LC/LB), and mid cognitive scores (MC/LB), respectively (Supplementary Figure 8B, 8C & 8D). The remaining profiles (C1, C2, C4, and C5) represent mid-cognitive scores with mid-stress/internalization scores (MC/MSI), high stress/internalization scores (MC/HSI), mid-behavioral scores (MC/MB), and high externalization score (MC/HE), respectively (Supplementary Figure 8B, 8C & 8D). Those results suggest that the extracted profiles within ABCD are consistent across multiple study populations. Using a one-way ANOVA and Tukey HSD pairwise comparison test, we evaluated the difference in raw cognitive and behavioral means across each profile (Supplementary Tables 12, 13, & 14).

| **Environmental Factors** | **Instrument name** | **Encoding method/Variables name** |
| --- | --- | --- |
| *Perinatal factors* | | |
| Maternal Age at delivery (years) | Developmental History (ph_p_dhx) | devhx_3_p |
| Gestational Age (weeks) |  | 40 if devhx_12a_p is 1, 40 -devhx_12_p if 0 |
| Birth Weight (kgs) |  | (birth_weight_lbs * 0.453592) + (birth_weight_oz * 0.0283495) |
| Substance Use (2, 1, or 0) |  | Sum of both parents (1 if 1 for devhx_8_tobacco, devhx_8_alcool, devhx_8_marijuana, devhx_8_coc_crack, devhx_8_her_morph, devhx_8_oxycont, or devhx_8_other_drugs) for father substance use, replace 8 by 9 in the variables name. |
| Total conditions (range from 0 to 13) |  | Sum of devhx_10a3_p, devhx_10b3_p, devhx_10c3_p, devhx_10d3_p, devhx_10e3_p, devhx_10f3_p, devhx_10g3_p, devhx_10h3_p, devhx_10i3_p, devhx_10j3_p, devhx_10k3_p, devhx_10l3_p,  devhx_10m3_p |
| Planned Pregnancy (1, 0) |  | devhx_6_p |
| *Adverse Childhood Experiences (ACEs)* | | |
| History of traumatic events (1, 0) | Parental KSADS – Symptoms and Diagnoses (mh_p_ksads_ss) | ksads_21_134_p |
| Family Conflict (range from 0 to 9) | Youth PhenX Family Environment Scale (ce_p_fes) | Total raw score (fes_y_ss_fc) |
| Parental Psychopathology (continuous score) | Adult Self Report (mh_p_asr) and Family History (mh_p_fhx) | Coded 1 if either of parents had history of fights, problems holding jobs, and with the police (famhx_ss_momdad_trb_p and famhx_ss_fath_prob_trb_p). Then, sum of the zscore of the total problem scale (asr_scr_totprob_t) and zscore of the new coded history variable. |
| *Sleep* | | |
| Sleeping Hours (1, 2, 3, 4, or 5) | Sleep Disturbance Scale for Children (ph_p_sds) | 1: 9-11h, 2: 8-9h, 3: 7-8h, 4:5-7h, 5: less than 5h (sleepdisturb1_p) |
| *Community/School Factors* | | |
| Neighborhood Safety (range from 1 to 5) | PhenX Neighborhood Safety/Crime Survey (ce_p_nsc) | Mean Safety score (nsc_p_ss_mean_3_items) |
| School environment (range from 5 to 24) | PhenX School Risk and Protective Factors (ce_y_srpf) | SRPF Environment subscale (srpf_y_ss_ses) |
| School involvement (range from 3 to 16) |  | SRPF Involvement subscale (srpf_y_ss_iiss) |
| School disengagement (range from 2 to 8) |  | SRPF Disengagement subscale (srpf_y_ss_dfs) |
| *Parental Factors* | | |
| Acceptance (range from 2 to 6) | Children’s Report of Parental Behavioral Inventory (ce_y_crpbi) | Mean Acceptance subscale from both parents (crpbi_y_ss_parent and crpbi_y_ss_caregiver) |
| Monitoring (range from 1 to 5) | Parental Monitoring (ce_y_pm) | Mean score (pmq_y_ss_mean) |
| Education Level (1, 2, 3, 4, or 5) | Demographics Questionnaire (abcd_p_demo) | Recoded: 1: No highschool, 2: highschool, GED, or equivalent, 3: Some college, 4: Bachelor, and 5: Postgraduate (Highest level from either parents) (demo_prnt_ed_v2, demo_prtnr_ed_v2) |
| *Economic Factors* | | |
| Ability to pay bills (1, 0) | Demographics Questionnaire (abcd_p_demo) | Coded 1 if went without telephone service or got services turned off (Gaz, electricity, etc.) from an inability to pay, else coded 0. (demo_fam_exp2_v2 and demo_fam_exp5_v2) |
| Ability to provide food (1, 0) |  | demo_fam_exp1_v2 |
| Ability to provide housing (1, 0) |  | Coded 1 if unable to pay the full amount of mortgage or facing eviction for an inability to pay, else coded 0. (demo_fam_exp3_v2 and demo_fam_exp4_v2) |
| Ability to provide medical care (1, 0) |  | Coded 1 if couldn’t go to the hospital/doctor/dentist because of an inability to pay, else coded 0. (demo_fam_exp6_v2 and demo_fam_exp7_v2) |

**Supplementary Table 1.** List of all measures and coding methods to extract environmental factors from the ABCD study.

|  | **Latent Factors** | | | **Communalities** |
| --- | --- | --- | --- | --- |
|  | **VA** | **EF/PS** | **MEM** |  |
| Eigenvalues | 3.47 | 1.22 | 0.96 | - |
| **Variables** | | | |  |
| PVT | **0.654** | -0.001 | 0.040 | 0.430 |
| FICA | 0.056 | **0.598** | -0.028 | 0.362 |
| LSWM | 0.340 | 0.088 | 0.301 | 0.214 |
| DCCS | 0.055 | **0.608** | 0.063 | 0.377 |
| PCPS | -0.086 | **0.649** | -0.025 | 0.429 |
| PSMT | -0.060 | 0.018 | **0.694** | 0.485 |
| ORR | **0.763** | 0.021 | -0.055 | 0.586 |
| RAVLT | 0.129 | 0.016 | **0.532** | 0.300 |
| LMT | 0.268 | 0.176 | 0.098 | 0.112 |
| Matrix | **0.405** | -0.010 | 0.221 | 0.213 |

**Supplementary Table 2.** Latent factor loadings for all variables and associated eigenvalues for the ABCD study during the baseline follow-up. Loadings > 0.40 are bolded. VA: Verbal Ability, EF/PS: Executive function and processing speed, and MEM: memory.

| **Latent Factors** | **Coefficients** | **Std. Error** | | **Z-value** | | **p-value** | |
| --- | --- | --- | --- | --- | --- | --- | --- |
| *Verbal Ability (VA)* | | | | | | | |
| ORR | 1 | | - | | - | | - |
| PVT | 0.938 | | 0.027 | | 34.48 | | < 0.001 |
| LSWM | 0.438 | | 0.033 | | 13.25 | | < 0.001 |
| *Executive Function/Processing Speed (EF/PS)* | | | | | | | |
| PCPS | 1 | | - | | - | | - |
| DCCS | 1.218 | | 0.039 | | 30.87 | | < 0.001 |
| FICA | 1.068 | | 0.036 | | 29.93 | | < 0.001 |
| *Memory (MEM)* | | | | | | | |
| PSMT | 1 | | - | | - | | - |
| RAVLT | 1.057 | | 0.039 | | 27.40 | | < 0.001 |
| LSWM | 0.582 | | 0.044 | | 13.27 | | < 0.001 |

**Supplementary Table 3.** ABCD baseline follow-up split-sample Confirmatory Factor Analysis (CFA) coefficients, standard errors, z-values, and p-values.

|  | **Latent Factors** | | | **Communalities** |
| --- | --- | --- | --- | --- |
|  | **VA** | **EF/PS** | **MEM** |  |
| Eigenvalues | 2.64 | 1.05 | 1.02 | - |
| **Variables** | | | |  |
| PVT | **0.693** | -0.008 | 0.058 | 0.483 |
| FICA | 0.136 | **0.491** | 0.019 | 0.260 |
| PCPS | -0.032 | **0.764** | -0.002 | 0.585 |
| PSMT | -0.039 | 0.011 | **0.722** | 0.523 |
| ORR | **0.790** | 0.011 | -0.034 | 0.625 |
| RAVLT | 0.131 | -0.015 | **0.497** | 0.264 |
| LMT | 0.233 | 0.115 | 0.215 | 0.114 |
| DICE | 0.152 | 0.044 | -0.001 | 0.025 |

**Supplementary Table 4.** Latent factor loadings for all variables and associated eigenvalues for the ABCD study during the 2-year follow-up. Loadings > 0.40 are bolded. VA: Verbal Ability, EF/PS: Executive function and processing speed, and MEM: memory.

| **Latent Factors** | **Coefficients** | **Std. Error** | | **Z-value** | | **p-value** | |
| --- | --- | --- | --- | --- | --- | --- | --- |
| *Verbal Ability (VA)* | | | | | | | |
| ORR | 1 | | - | | - | | - |
| PVT | 1.036 | | 0.047 | | 22.06 | | < 0.001 |
| *Executive Function/Processing Speed (EF/PS)* | | | | | | | |
| PCPS | 1 | | - | | - | | - |
| FICA | 1.047 | | 0.074 | | 14.18 | | < 0.001 |
| *Memory (MEM)* | | | | | | | |
| PSMT | 1 | | - | | - | | - |
| RAVLT | 1.014 | | 0.064 | | 15.86 | | < 0.001 |

**Supplementary Table 5.** ABCD 2-year follow-up split-sample Confirmatory Factor Analysis (CFA) coefficients, standard errors, z-values, and p-values.

|  | **Latent Factors** | | | **Communalities** |
| --- | --- | --- | --- | --- |
|  | **VA** | **EF/PS** | **MEM** |  |
| Eigenvalues | 2.88 | 1.12 | 0.97 | - |
| **Variables** | | | |  |
| PVT | **0.677** | -0.051 | 0.156 | 0.486 |
| FICA | 0.065 | **0.694** | -0.023 | 0.486 |
| LSWM | 0.200 | 0.029 | **0.503** | 0.294 |
| PCPS | -0.060 | **0.680** | 0.050 | 0.468 |
| PSMT | -0.061 | 0.072 | **0.576** | 0.341 |
| ORR | **0.808** | 0.051 | -0.067 | 0.661 |
| LMT | 0.230 | 0.204 | 0.141 | 0.114 |
| DICE | -0.190 | -0.011 | -0.125 | 0.052 |
| BIRD | 0.056 | 0.057 | 0.130 | 0.023 |

**Supplementary Table 6.** Latent factor loadings for all variables and associated eigenvalues for the ABCD study during the 4-year follow-up. Loadings > 0.40 are bolded. VA: Verbal Ability, EF/PS: Executive function and processing speed, and MEM: memory.

| **Latent Factors** | **Coefficients** | **Std. Error** | | **Z-value** | | **p-value** | |
| --- | --- | --- | --- | --- | --- | --- | --- |
| *Verbal Ability (VA)* | | | | | | | |
| ORR | 1 | | - | | - | | - |
| PVT | 0.959 | | 0.049 | | 19.49 | | < 0.001 |
| *Executive Function/Processing Speed (EF/PS)* | | | | | | | |
| PCPS | 1 | | - | | - | | - |
| FICA | 1.266 | | 0.140 | | 9.02 | | < 0.001 |
| *Memory (MEM)* | | | | | | | |
| PSMT | 1 | | - | | - | | - |
| LSWM | 1.723 | | 0.157 | | 10.97 | | < 0.001 |

**Supplementary Table 7.** ABCD 4-year follow-up split-sample Confirmatory Factor Analysis (CFA) coefficients, standard errors, z-values, and p-values.

|  | **Latent Factors** | | | **Communalities** |
| --- | --- | --- | --- | --- |
|  | **VA** | **EF/PS** | **MEM** |  |
| Eigenvalues | 3.20 | 1.32 | 0.93 | - |
| **Variables** | | | |  |
| DCCS | 0.193 | **0.481** | 0.089 | 0.277 |
| FICA | -0.034 | **0.913** | 0.034 | 0.836 |
| LSWM | **0.632** | -0.015 | 0.271 | 0.473 |
| ORR | **0.624** | 0.020 | -0.072 | 0.394 |
| PCPS | 0.123 | **0.485** | -0.258 | 0.317 |
| PennWM | 0.046 | 0.154 | **0.525** | 0.301 |
| PennMR | **0.457** | 0.014 | **0.368** | 0.345 |
| WASI-VC | **0.826** | 0.039 | -0.091 | 0.692 |

**Supplementary Table 8.** Latent factors loadings and eigenvalues for the BANDA study. Loadings > 0.40 are bolded. VA: Verbal Ability, EF/PS: Executive Function/Processing Speed, and MEM: Memory.

|  | **Coefficients** | **Std. Error** | **Z-value** | **p-value** |
| --- | --- | --- | --- | --- |
| **Latent Factors** | | | | |
| *Verbal Ability (VA)* | | | | |
| ORR | 1 | - | - | - |
| WASI-VC | 1.171 | 0.263 | 4.45 | < 0.001 |
| LSWM | 0.387 | 0.183 | 2.12 | 0.034 |
| *Executive Function/Processing Speed (EF/PS)* | | | | |
| DCCS | 1 | - | - | - |
| FICA | 1.297 | 0.313 | 4.14 | < 0.001 |
| PCPS | 0.965 | 0.228 | 4.23 | < 0.001 |
| *Memory (MEM)* | | | | |
| PennWM | 1.000 | - | - | - |
| PennMR | 3.387 | 3.000 | 1.13 | 0.259 |
| LSWM | 0.440 | 0.417 | 1.05 | 0.292 |

**Supplementary Table 9.** BANDA Split-sample Confirmatory Factor Analysis (CFA) coefficients, standard errors, z-values, and p-values.

|  | **Latent Factors** | | | **Communalities** |
| --- | --- | --- | --- | --- |
|  | **VA** | **EF/PS** | **MEM** |  |
| Eigenvalues | 2.17 | 1.19 | 1.03 | - |
| **Variables** | | | |  |
| BL | 0.005 | 0.072 | **0.593** | 0.357 |
| SI | **0.510** | -0.045 | 0.333 | 0.373 |
| MR | -0.081 | 0.258 | 0.366 | 0.207 |
| DS | 0.037 | 0.157 | 0.038 | 0.028 |
| CO | -0.021 | -0.115 | 0.396 | 0.170 |
| VC | **1.003** | 0.018 | -0.033 | 1.008 |
| BA | 0.009 | **0.996** | 0.003 | 0.992 |

**Supplementary Table 10.** Latent factors loadings and eigenvalues for the GESTE study. Loadings > 0.40 are bolded. VA: Verbal Ability, MEM: Memory, and EF/PS: Executive Function/Processing Speed.

|  | **Coefficients** | **Std. Error** | **Z-value** | **p-value** |
| --- | --- | --- | --- | --- |
| **Latent Factors** | | | | |
| *Verbal Ability (VA)* | | | | |
| VC | 1 | - | - | - |
| SI | 0.941 | 0.196 | 4.79 | < 0.001 |
| *Executive Function/Processing Speed (EF/PS)* | | | | |
| CO | 1.000 | - | - | - |
| *Memory (MEM)* | | | | |
| BL | 1 | - | - | - |
| MR | 0.725 | 0.132 | 5.48 | < 0.001 |
| DS | 0.718 | 0.138 | 5.20 | < 0.001 |
| BA | 1.051 | 0.154 | 6.84 | < 0.001 |

**Supplementary Table 11.** GESTE Split-sample Confirmatory Factor Analysis (CFA) coefficients, standard errors, z-values, and p-values.

|  | **2-year follow-up** | | **4-year follow-up** | |
| --- | --- | --- | --- | --- |
|  | **Male** | **Female** | **Male** | **Female** |
| **N (%)** | 3839 (52.1) | 3529 (47.89) | 1481 (52.04) | 1364 (47.93) |
| **Age (std)** | 11.97 (0.65) | 11.94 (0.65) | 14.08 (0.71) | 14.08 (0.69) |
| **Race/Ethnicity, count (%)** | | | | |
| White | 2176, (29.53) | 1948 (26.44) | 841 (29.55) | 743 (26.11) |
| Black or African American | 450 (6.11) | 455 (6.17) | 146 (5.13) | 162 (5.69) |
| Hispanic or Latino | 740 (10.04) | 679 (9.21) | 320 (11.24) | 269 (9.45) |
| Asian | 80 (1.09) | 77 (1.04) | 34 (1.19) | 31 (1.09) |
| Other | 393 (5.33) | 370 (5.02) | 140 (4.92) | 159 (5.59) |
| **Highest parental education, count (%)** | | | | |
| No Highschool | 161 (2.18) | 158 (2.14) | 63 (2.21) | 65 (2.28) |
| Highschool, GED, or equivalent | 319 (4.33) | 277 (3.76) | 108 (3.79) | 93 (3.27) |
| Some college | 951 (12.91) | 878 (11.91) | 373 (13.11) | 339 (11.91) |
| Bachelor Degree | 1055 (14.32) | 949 (12.88) | 414 (14.55) | 367 (12.9) |
| Postgraduate Degree | 1353 (18.36) | 1267 (17.19) | 523 (18.38) | 500 (17.57) |
| **Familial Income (USD$), count (%)** | | | | |
| < 50 000$USD | 660 (8.96) | 596 (8.09) | 250 (8.78) | 215 (7.55) |
| 50 000-100 000$USD | 1326 (17.99) | 1276 (17.32) | 515 (18.1) | 495 (17.39) |
| > 100 000$USD | 1554 (21.09) | 1399 (18.98) | 606 (21.29) | 567 (19.92) |
| **Cognitive and Behavioral Symptoms Scores, mean (std) ^φ^** | | | | |
| Internalization | -0.01 (0.94) | 0.01 (1.01) | -0.0 (0.86) | 0.0 (1.1) |
| Externalization | 0.0 (1.01) | 0.0 (0.91) | 0.01 (1.0) | -0.0 (0.89) |
| Stress | 0.0 (0.97) | 0.0 (0.96) | 0.0 (0.94) | -0.0 (1.05) |
| VA | -0.02 (0.60) | 0.02 (0.6) | -0.01 (0.66) | 0.01 (0.67) |
| EFPS | -0.01 (0.49) | 0.01 (0.48) | -0.01 (0.48) | 0.01 (0.48) |
| MEM | -0.01 (0.45) | 0.01 (0.44) | -0.01 (0.35) | 0.01 (0.35) |

**Supplementary Table 12.** The demographic table for the ABCD 2-year and 4-year follow-up included participants. GED: General Equivalent Diploma. **^φ^**: The mean and std values presented were computed after harmonization.

| **Variables** | **ABCD Baseline follow-up** | | | | | | | |
| --- | --- | --- | --- | --- | --- | --- | --- | --- |
|  | **F** | ***p*** | **P1 vs P2** | **P1 vs P3** | **P1 vs P4** | **P2 vs P3** | **P2 vs P4** | **P3 vs P4** |
| **Internalization** | 4040.00 | < 0.001 | < 0.001 | < 0.001 | < 0.001 | < 0.001 | < 0.001 | 0.013 |
| **Externalization** | 4560.57 | < 0.001 | < 0.001 | < 0.001 | < 0.001 | < 0.001 | < 0.001 | 0.180 |
| **Stress** | 3641.73 | < 0.001 | < 0.001 | < 0.001 | < 0.001 | < 0.001 | < 0.001 | < 0.001 |
| **VA** | 2595.80 | < 0.001 | < 0.001 | < 0.001 | < 0.001 | < 0.001 | < 0.001 | < 0.001 |
| **EFPS** | 1873.11 | < 0.001 | < 0.001 | < 0.001 | < 0.001 | < 0.001 | < 0.001 | < 0.001 |
| **MEM** | 2583.37 | < 0.001 | < 0.001 | < 0.001 | < 0.001 | < 0.001 | < 0.001 | < 0.001 |

**Supplementary Table 13.** One-way ANOVA statistical results between ABCD baseline follow-up profiles for each cognitive and behavioral variable with post-hoc Tukey HSD analysis for pairwise comparison of each profile (the value represents the p-value). F: F-statistic, p: p-value. VA: Verbal Ability, EFPS: Executive Function/Processing Speed, and MEM: Memory. P1: Profile 1 (MC/HSI), P2: Profile 2 (MC/HE), P3: Profile 3 (HC/LB), P4: Profile 4 (LC/LB).

| **Variables** | **BANDA** | | | | | | | |
| --- | --- | --- | --- | --- | --- | --- | --- | --- |
|  | **F** | ***p*** | **P1 vs P2** | **P1 vs P3** | **P1 vs P4** | **P2 vs P3** | **P2 vs P4** | **P3 vs P4** |
| **Internalization** | 89.34 | < 0.001 | < 0.001 | < 0.001 | < 0.001 | < 0.001 | < 0.001 | 0.124 |
| **Externalization** | 107.10 | < 0.001 | < 0.001 | < 0.001 | < 0.001 | < 0.001 | < 0.001 | 0.935 |
| **Stress** | 91.67 | < 0.001 | 0.428 | < 0.001 | < 0.001 | < 0.001 | < 0.001 | 0.265 |
| **VA** | 35.14 | < 0.001 | 0.496 | < 0.001 | < 0.001 | < 0.001 | 0.002 | < 0.001 |
| **EFPS** | 14.83 | < 0.001 | 0.999 | 0.001 | 0.065 | 0.006 | 0.079 | < 0.001 |
| **MEM** | 21.61 | < 0.001 | 0.894 | 0.001 | 0.001 | < 0.001 | 0.034 | < 0.001 |

**Supplementary Table 14.** One-way ANOVA statistical results between BANDA profiles for each cognitive and behavioral variable with post-hoc Tukey HSD analysis for pairwise comparison of each profile (the value represents the p-value). F: F-statistic, p: p-value. VA: Verbal Ability, EFPS: Executive Function/Processing Speed, and MEM: Memory. P1: Profile 1 (MC/HSI), P2: Profile 2 (MC/HE), P3: Profile 3 (HC/LB), P4: Profile 4 (LC/LB).

| **Variables** | **GESTE** | | | | | | | |
| --- | --- | --- | --- | --- | --- | --- | --- | --- |
|  | **F** | ***p*** | **P1 vs P2** | **P1 vs P3** | **P1 vs P4** | **P2 vs P3** | **P2 vs P4** | **P3 vs P4** |
| **Internalization** | 103.44 | < 0.001 | < 0.001 | < 0.001 | < 0.001 | < 0.001 | < 0.001 | 0.893 |
| **Externalization** | 98.94 | < 0.001 | < 0.001 | < 0.001 | < 0.001 | < 0.001 | < 0.001 | 0.966 |
| **Stress** | 108.42 | < 0.001 | 0.733 | < 0.001 | < 0.001 | < 0.001 | < 0.001 | 0.988 |
| **VA** | 50.52 | < 0.001 | 0.607 | < 0.001 | < 0.001 | < 0.001 | 0.002 | < 0.001 |
| **EFPS** | 38.46 | < 0.001 | 0.883 | < 0.001 | 0.007 | < 0.001 | < 0.001 | < 0.001 |
| **MEM** | 43.25 | < 0.001 | 0.950 | < 0.001 | 0.001 | < 0.001 | < 0.001 | < 0.001 |

**Supplementary Table 15.** One-way ANOVA statistical results between GESTE profiles for each cognitive and behavioral variable with post-hoc Tukey HSD analysis for pairwise comparison of each profile (the value represents the p-value). F: F-statistic, p: p-value. VA: Verbal Ability, EFPS: Executive Function/Processing Speed, and MEM: Memory. P1: Profile 1 (MC/HSI), P2: Profile 2 (MC/HE), P3: Profile 3 (HC/LB), P4: Profile 4 (LC/LB).

| **Variables** | **ABCD 2-year follow-up** | | | | | | | |
| --- | --- | --- | --- | --- | --- | --- | --- | --- |
|  | **F** | ***p*** | **P1 vs P2** | **P1 vs P3** | **P1 vs P4** | **P2 vs P3** | **P2 vs P4** | **P3 vs P4** |
| **Internalization** | 2777.66 | < 0.001 | < 0.001 | < 0.001 | < 0.001 | < 0.001 | < 0.001 | 0.003 |
| **Externalization** | 3046.96 | < 0.001 | < 0.001 | < 0.001 | < 0.001 | < 0.001 | < 0.001 | 0.940 |
| **Stress** | 2611.17 | < 0.001 | < 0.001 | < 0.001 | < 0.001 | < 0.001 | < 0.001 | 0.333 |
| **VA** | 1848.32 | < 0.001 | < 0.001 | < 0.001 | < 0.001 | < 0.001 | < 0.001 | < 0.001 |
| **EFPS** | 1041.82 | < 0.001 | 0.755 | < 0.001 | < 0.001 | < 0.001 | < 0.001 | < 0.001 |
| **MEM** | 1446.36 | < 0.001 | < 0.001 | < 0.001 | < 0.001 | < 0.001 | < 0.001 | < 0.001 |

**Supplementary Table 16.** One-way ANOVA statistical results between ABCD 2-year follow-up profiles for each cognitive and behavioral variable with post-hoc Tukey HSD analysis for pairwise comparison of each profile (the value represents the p-value). F: F-statistic, p: p-value. VA: Verbal Ability, EFPS: Executive Function/Processing Speed, and MEM: Memory. P1: Profile 1 (MC/HSI), P2: Profile 2 (MC/HE), P3: Profile 3 (HC/LB), P4: Profile 4 (LC/LB).

| **Variables** | **ABCD 4-year follow-up** | | | | | | | |
| --- | --- | --- | --- | --- | --- | --- | --- | --- |
|  | **F** | ***p*** | **P1 vs P2** | **P1 vs P3** | **P1 vs P4** | **P2 vs P3** | **P2 vs P4** | **P3 vs P4** |
| **Internalization** | 1192.07 | < 0.001 | < 0.001 | < 0.001 | < 0.001 | < 0.001 | < 0.001 | 0.088 |
| **Externalization** | 1472.99 | < 0.001 | < 0.001 | < 0.001 | < 0.001 | < 0.001 | < 0.001 | 0.998 |
| **Stress** | 1089.46 | < 0.001 | 0.002 | < 0.001 | < 0.001 | < 0.001 | < 0.001 | 0.998 |
| **VA** | 790.72 | < 0.001 | < 0.001 | < 0.001 | < 0.001 | < 0.001 | < 0.001 | < 0.001 |
| **EFPS** | 348.11 | < 0.001 | 0.058 | < 0.001 | < 0.001 | < 0.001 | < 0.001 | < 0.001 |
| **MEM** | 615.41 | < 0.001 | < 0.001 | < 0.001 | < 0.001 | < 0.001 | < 0.001 | < 0.001 |

**Supplementary Table 17.** One-way ANOVA statistical results between ABCD 4-year follow-up profiles for each cognitive and behavioral variable with post-hoc Tukey HSD analysis for pairwise comparison of each profile (the value represents the p-value). F: F-statistic, p: p-value. VA: Verbal Ability, EFPS: Executive Function/Processing Speed, and MEM: Memory. P1: Profile 1 (MC/HSI), P2: Profile 2 (MC/HE), P3: Profile 3 (HC/LB), P4: Profile 4 (LC/LB).

| **Variables** | **F** | ***p*** | **ABCD vs BANDA (*p*)** | **ABCD vs GESTE (*p*)** | **BANDA vs GESTE (*p*)** |
| --- | --- | --- | --- | --- | --- |
| **Profile 1 (MC/HSI)** | | | | | |
| **Internalization** | 3.66 | **0.026*** | 0.062 | 0.266 | 0.797 |
| **Externalization** | 1.75 | 0.174 | 0.305 | 0.457 | 0.953 |
| **Stress** | 3.86 | **0.021*** | 0.567 | **0.025*** | 0.570 |
| **VA** | 0.28 | 0.754 | 0.845 | 0.860 | 0.998 |
| **EFPS** | 0.52 | 0.595 | 0.923 | 0.606 | 0.931 |
| **MEM** | 0.46 | 0.634 | 0.983 | 0.620 | 0.723 |
| **Profile 2 (MC/HE)** | | | | | |
| **Internalization** | 0.58 | 0.559 | 0.899 | 0.595 | 0.599 |
| **Externalization** | 2.09 | 0.124 | 0.942 | 0.106 | 0.559 |
| **Stress** | 1.78 | 0.169 | 0.681 | 0.201 | 0.903 |
| **VA** | 0.63 | 0.531 | 0.893 | 0.551 | 0.950 |
| **EFPS** | 0.56 | 0.570 | 0.828 | 0.641 | 0.994 |
| **MEM** | 1.18 | 0.307 | 0.793 | 0.335 | 0.922 |
| **Profile 3 (HC/LB)** | | | | | |
| **Internalization** | 17.21 | **< 0.001*** | **< 0.001*** | 0.187 | **0.006*** |
| **Externalization** | 2.90 | 0.060 | 0.065 | 0.627 | 0.523 |
| **Stress** | 11.03 | **< 0.001*** | **< 0.001*** | 0.705 | **< 0.001*** |
| **VA** | 0.10 | 0.906 | 0.966 | 0.930 | 0.903 |
| **EFPS** | 1.23 | 0.293 | 0.434 | 0.610 | 0.261 |
| **MEM** | 2.31 | 0.099 | 0.095 | 0.858 | 0.132 |
| **Profile 4 (LC/LB)** | | | | | |
| **Internalization** | 2.92 | 0.054 | 0.983 | **0.042*** | 0.339 |
| **Externalization** | 4.41 | **0.012*** | 0.363 | **0.021*** | 0.798 |
| **Stress** | 1.04 | 0.355 | 0.321 | 1.000 | 0.510 |
| **VA** | 0.09 | 0.910 | 0.906 | 0.996 | 0.920 |
| **EFPS** | 1.28 | 0.279 | 0.365 | 0.657 | 0.871 |
| **MEM** | 1.83 | 0.160 | 0.257 | 0.501 | 0.865 |

**Supplementary Table 18.** One-way ANOVA statistical results between the mean score for each variable between each cohort and across all profiles. Post hoc Tukey HSD pairwise comparison results between groups, values represent the p-values. F: F-statistic, p: p-value. VA: Verbal Ability, EFPS: Executive Function/Processing Speed, and MEM: Memory.

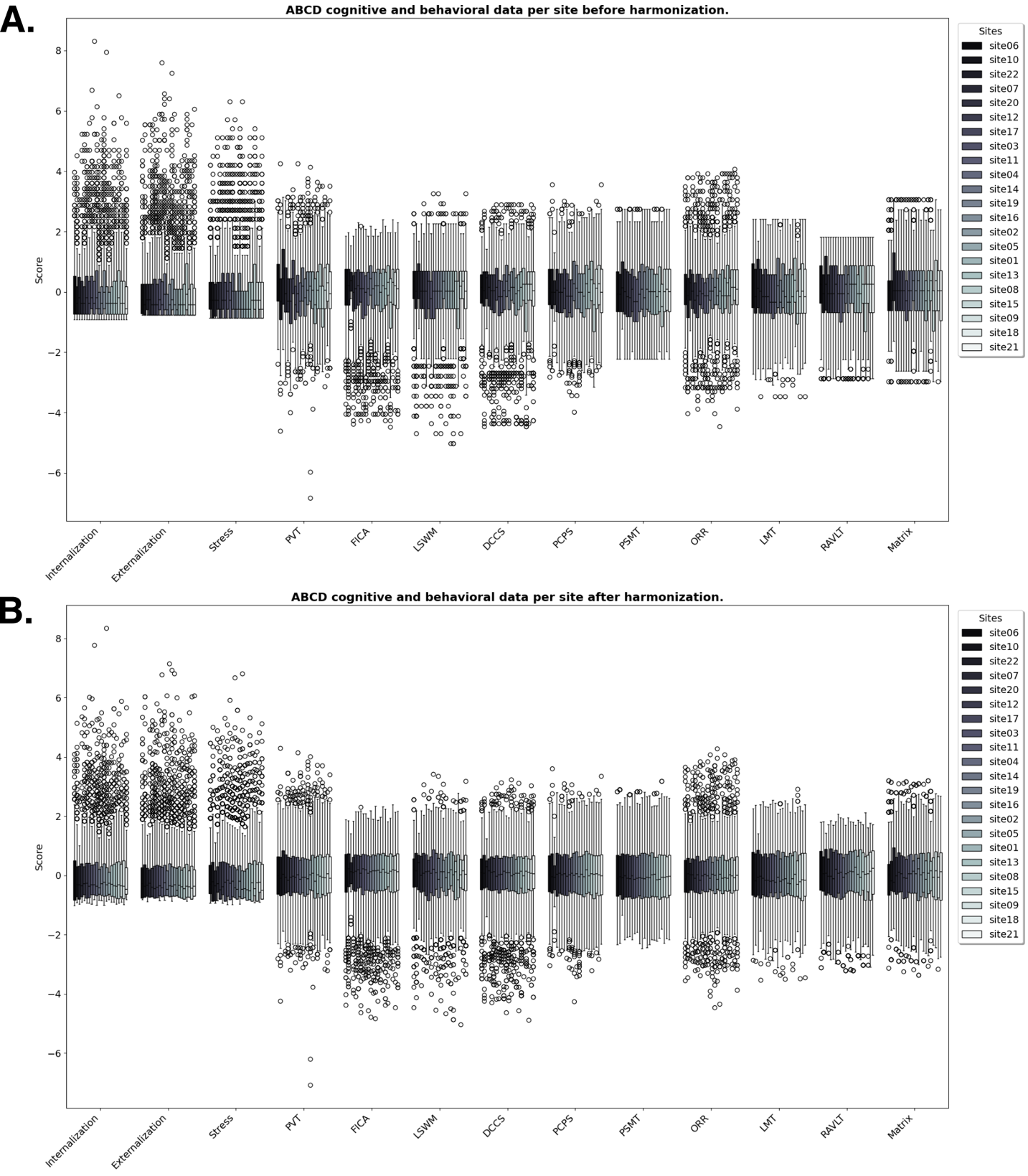

**Supplementary Figure 1.** Boxplot of the ABCD baseline follow-up site’s distributions for each cognitive and behavioral variable. **A.** Before harmonization. **B.** After harmonization. PVT: Picture Vocabulary Task, FICA: Flanker Inhibitory Control & Attention, LSWM: List Sorting Working Memory, DCCS: Dimensional Change Card Sorting, PCPS: Pattern Comparison Processing Speed, PSMT: Pattern Sequence Memory Task, ORR: Oral Reading Recognition, LMT: Little Man’s Task, RAVLT: Rey Auditory Verbal Learning Task, and Matrix: WISC-V Matrix Reasoning.

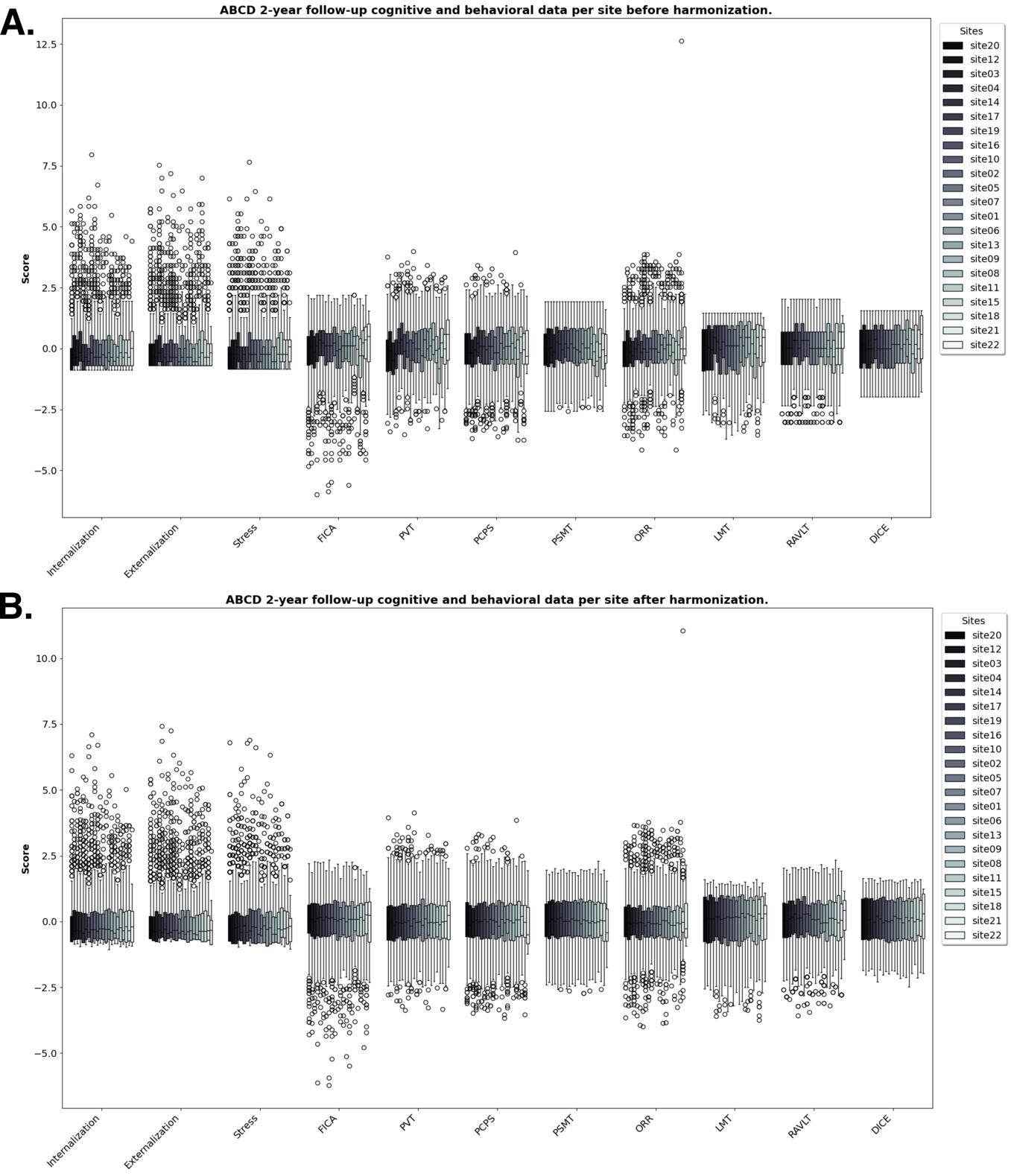

**Supplementary Figure 2.** Boxplot of the ABCD 2-year follow-up site’s distribution for each cognitive and behavioral variable. **A.** Before harmonization. **B.** After harmonization. PVT: Picture Vocabulary Task, FICA: Flanker Inhibitory Control & Attention, PCPS: Pattern Comparison Processing Speed, PSMT: Pattern Sequence Memory Task, ORR: Oral Reading Recognition, LMT: Little Man’s Task, RAVLT: Rey Auditory Verbal Learning Task, and DICE: Game of Dice Task.

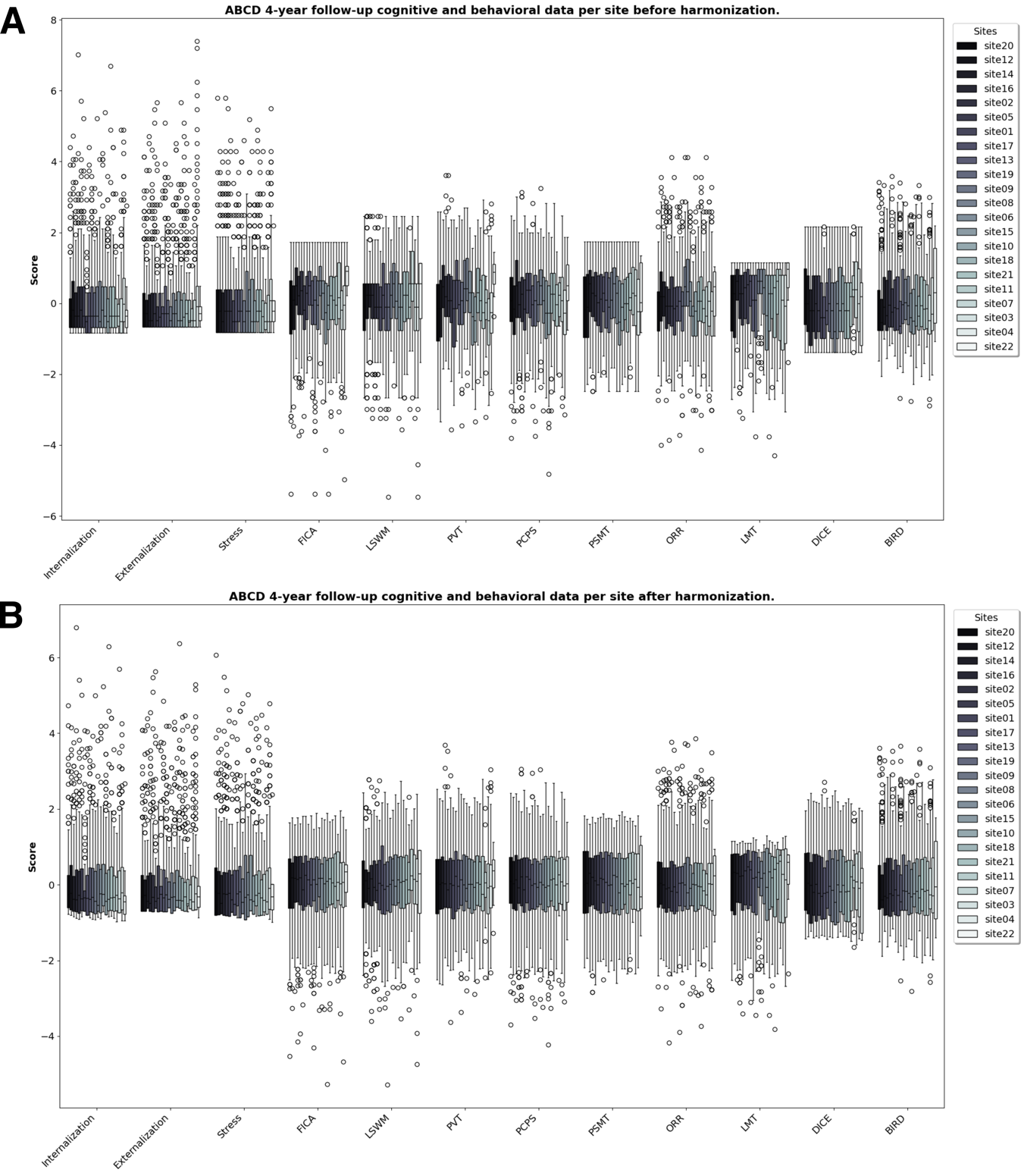

**Supplementary Figure 3.** Boxplot of the ABCD 4-year follow-up site’s distribution for each cognitive and behavioral variable. **A.** Before harmonization. **B.** After harmonization. PVT: Picture Vocabulary Task, LSWM: List Sorting Working Memory, FICA: Flanker Inhibitory Control & Attention, PCPS: Pattern Comparison Processing Speed, PSMT: Pattern Sequence Memory Task, ORR: Oral Reading Recognition, LMT: Little Man’s Task, DICE: Game of Dice Task, and BIRD: Behavioral Indicatory of Resiliency to Distress.

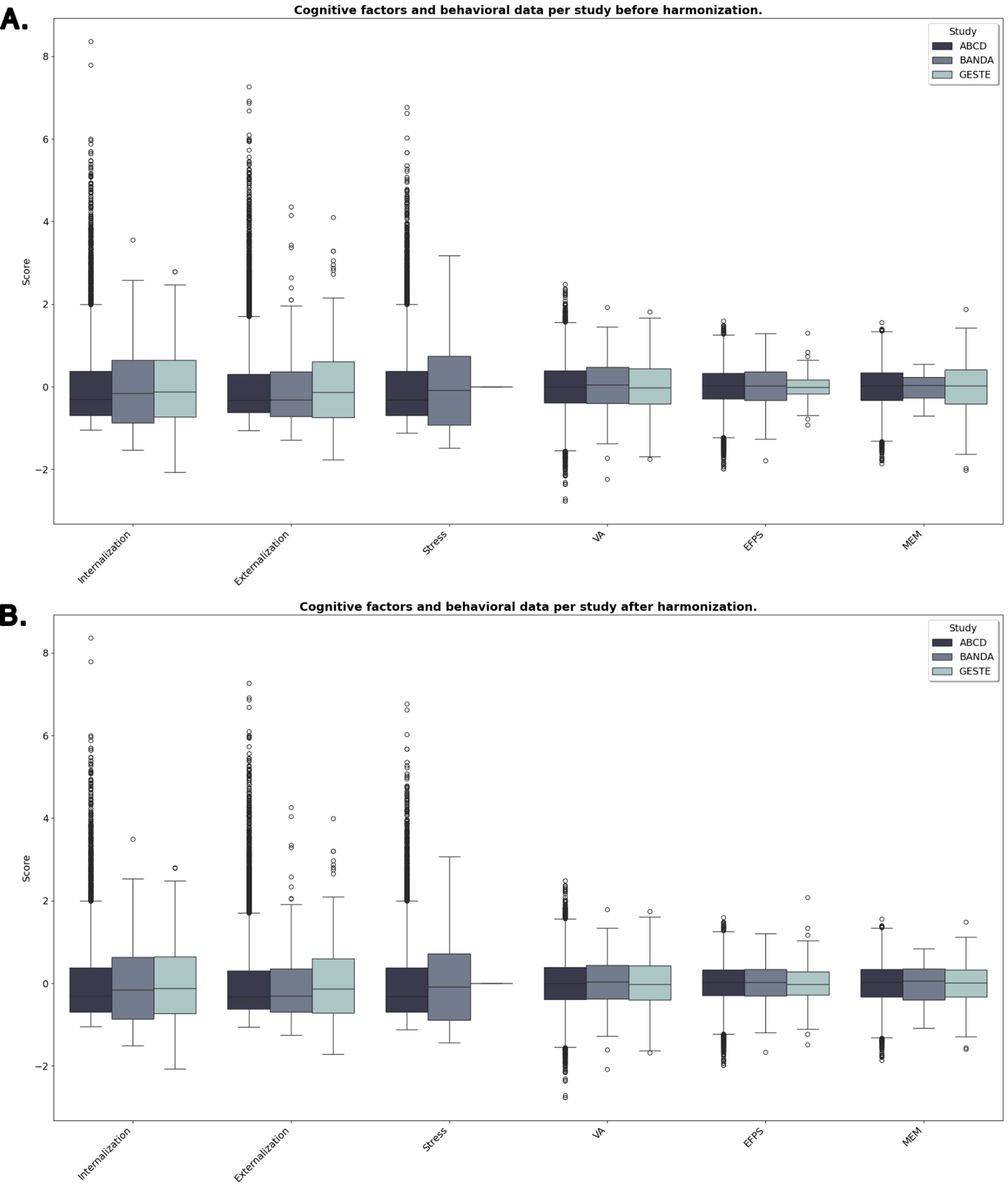

**Supplementary Figure 4.** Boxplot of the common variables distribution between the ABCD, GESTE, and BANDA study before and after harmonization using the ComBat framework. **A.** Before harmonization. **B.** After harmonization. VA: Verbal Ability, EF/PS: Executive Functions/Processing Speed, and MEM: Memory.

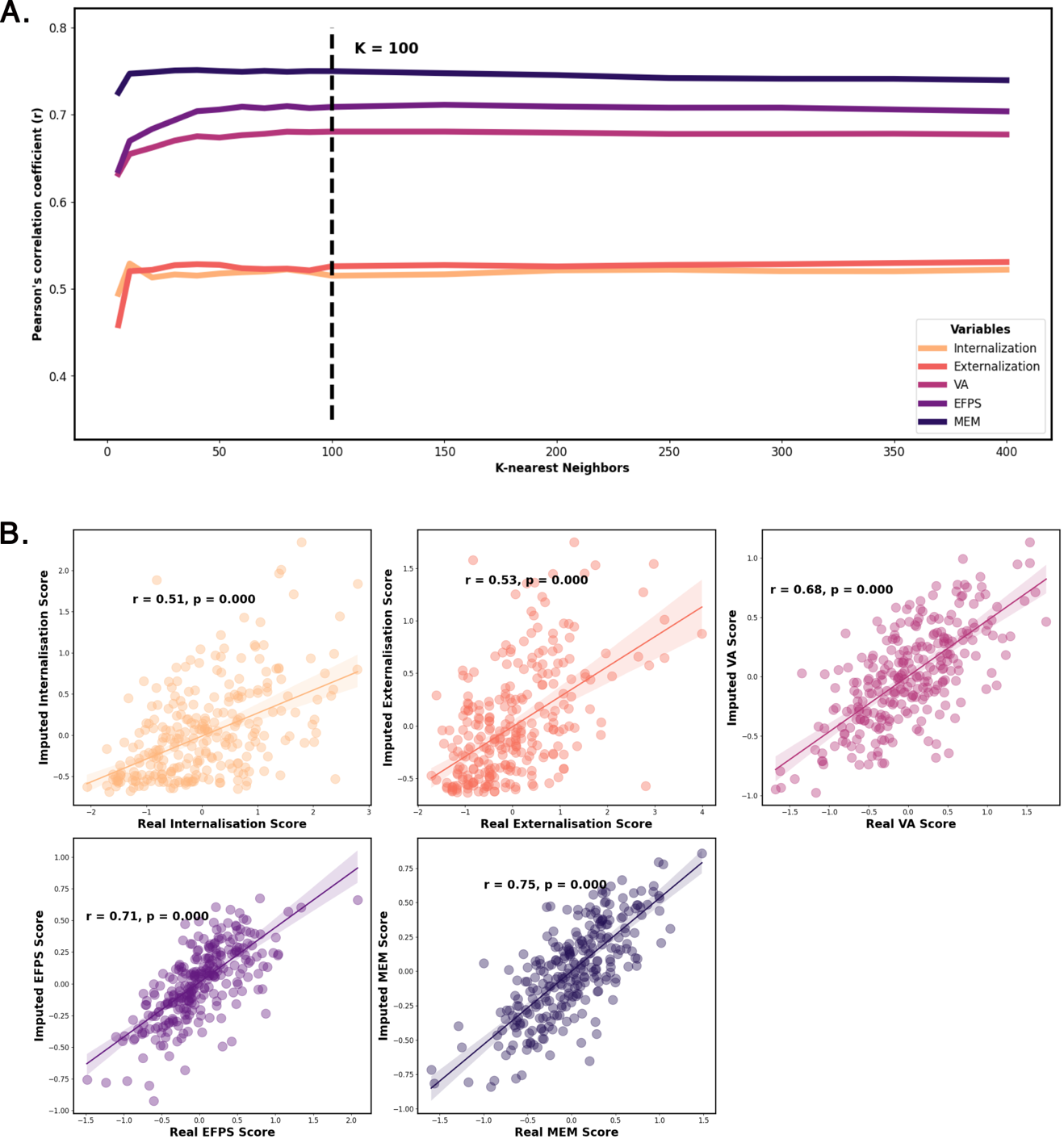

**Supplementary Figure 5.** Evaluation of the performance of the k-Nearest Neighbors imputation (KNN) model using the Euclidean distance as weight. **A.** Line plot showing the progression of Pearson’s correlation coefficient with the increase in the number of neighbors used when imputing GESTE variables with known real values. **B.** Correlation between the imputed values and the real values in the GESTE cohort using the optimal number neighbors (k = 100). r: Pearson’s correlation coefficient. VA: Verbal Ability, EFPS: Executive Function/Processing Speed, and MEM: Memory.

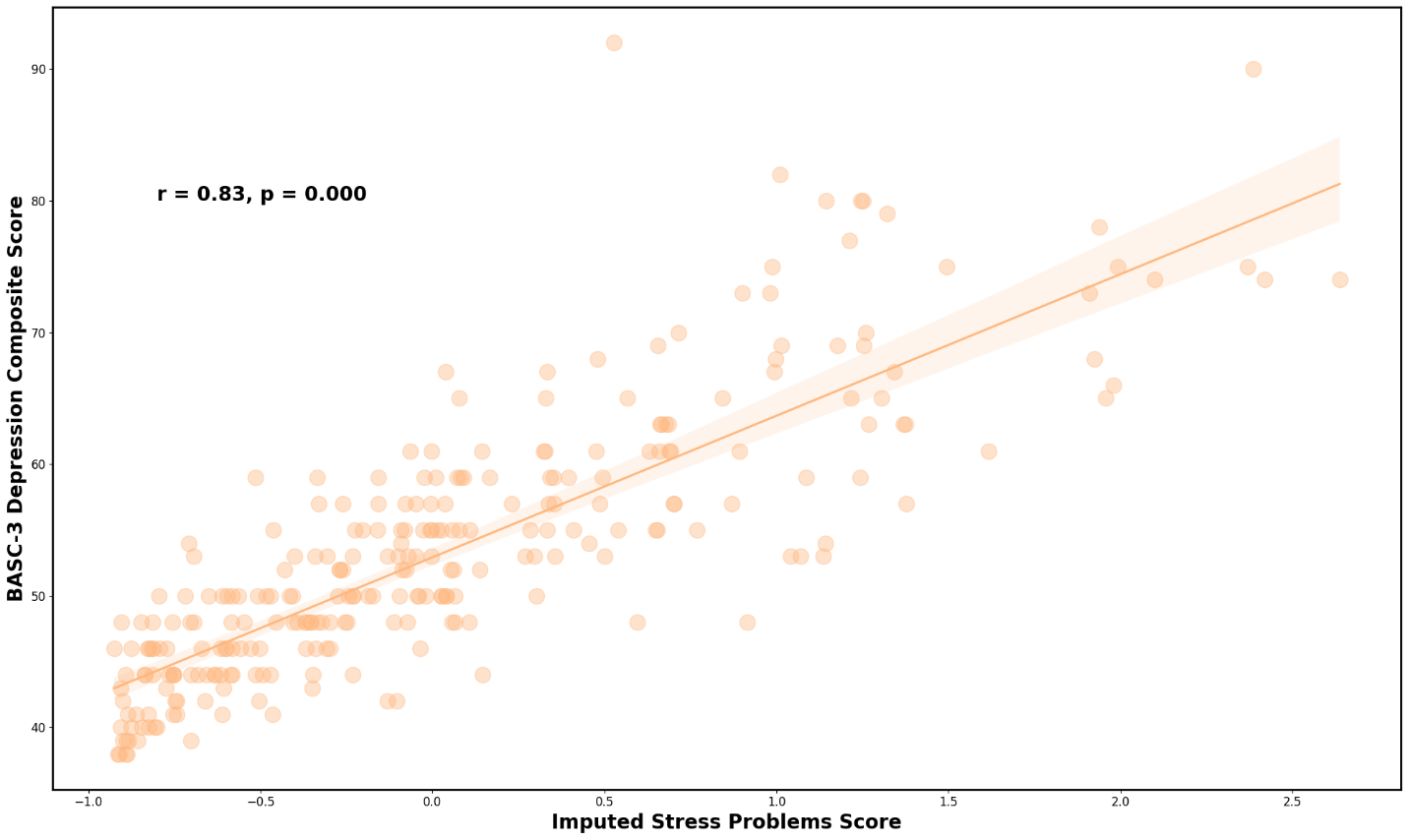

**Supplementary Figure 6.** Scatter plot of the imputed stress problems score compared to the BASC-3 Depression composite score reflecting stress and general unhappiness and sadness. r: Pearson’s correlation coefficient. BASC-3: Behavioral Assessment System for Children 3^rd^ edition.

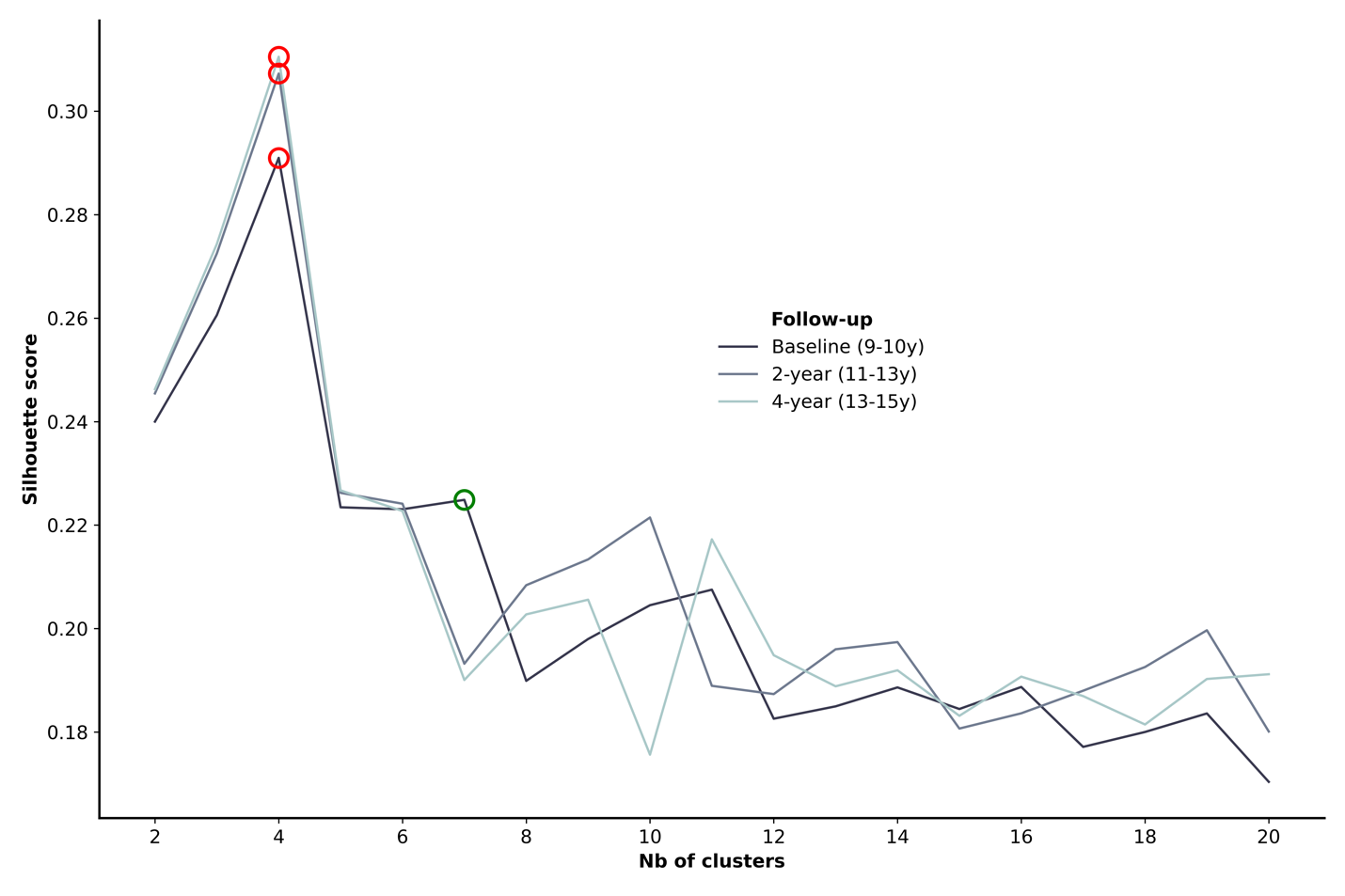

**Supplementary Figure 7.** Line plot of the silhouette score for each number of clusters tested. Each line represents a different follow-up within the ABCD cohort. The red circle represents the optimal number of cluster (derived by taking the highest silhouette score). The green circle represents a local minimum that could be selected for a more granular clustering.

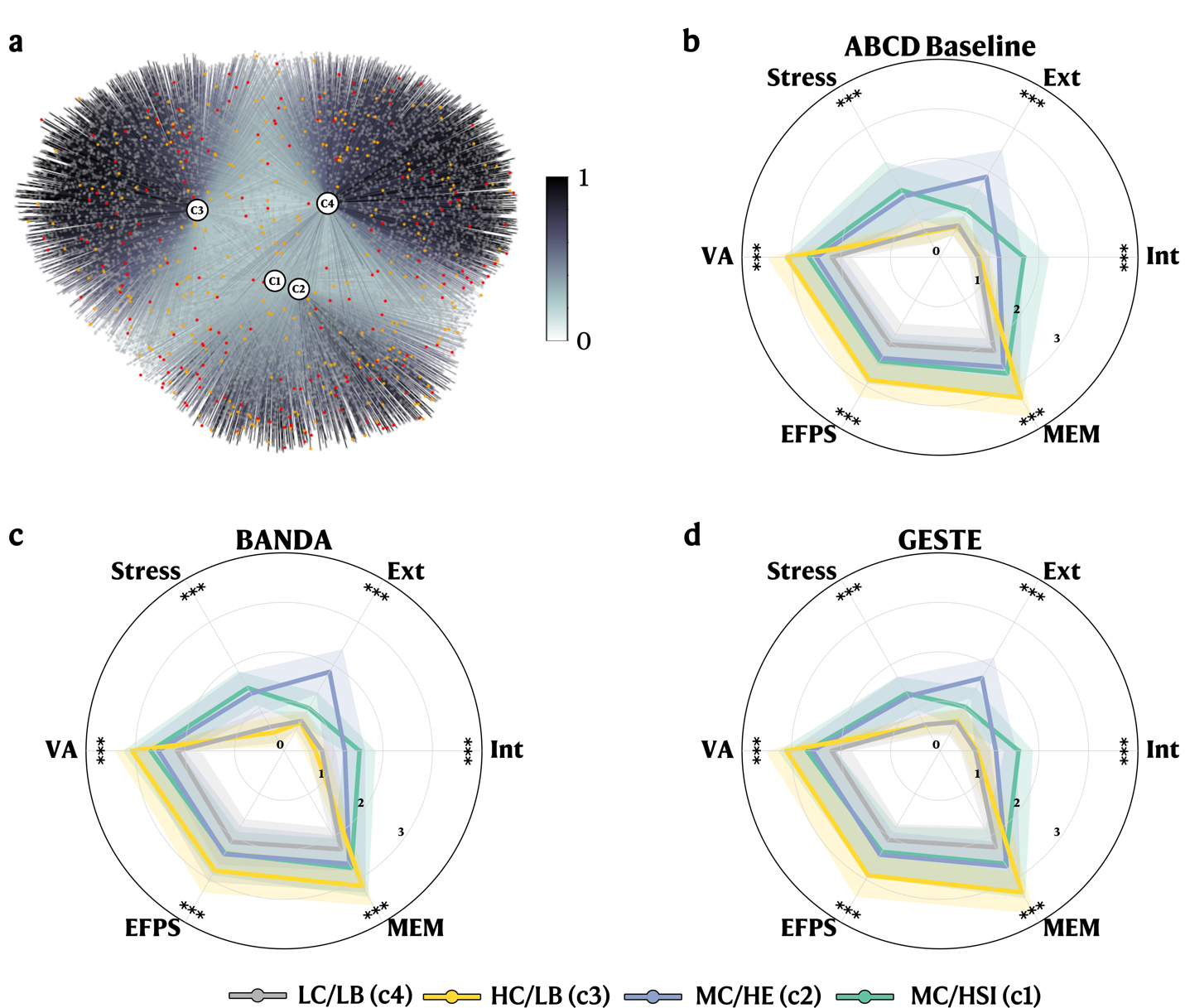

**Supplementary Figure 8.** FCM clustering results with radar plot for each study. FCM was initially computed on the ABCD dataset. The resulting centroids were used to predict the membership values for the BANDA and GESTE datasets. **A.** Graph Network including participants from all studies (Grey nodes: ABCD participants, red nodes: BANDA participants, and orange nodes: GESTE participants). Edges represent membership values to each cluster. **B.** Radar plot showing the mean values (with standard deviation) for each cognitive and behavioral variable stratified by clusters for the ABCD participants. **C.** Radar plot showing the mean values (with standard deviation) for each cognitive and behavioral variable stratified by clusters for the BANDA participants. **D.** Radar plot showing the mean values (with standard deviation) for each cognitive and behavioral variable stratified by clusters for the GESTE participants. ***: p < 0.001. Scores are scaled for visualization purposes. VA: Verbal Ability. EFPS: Executive Function and Processing Speed. MEM: Memory. Ext: Externalization. Int: Internalization.

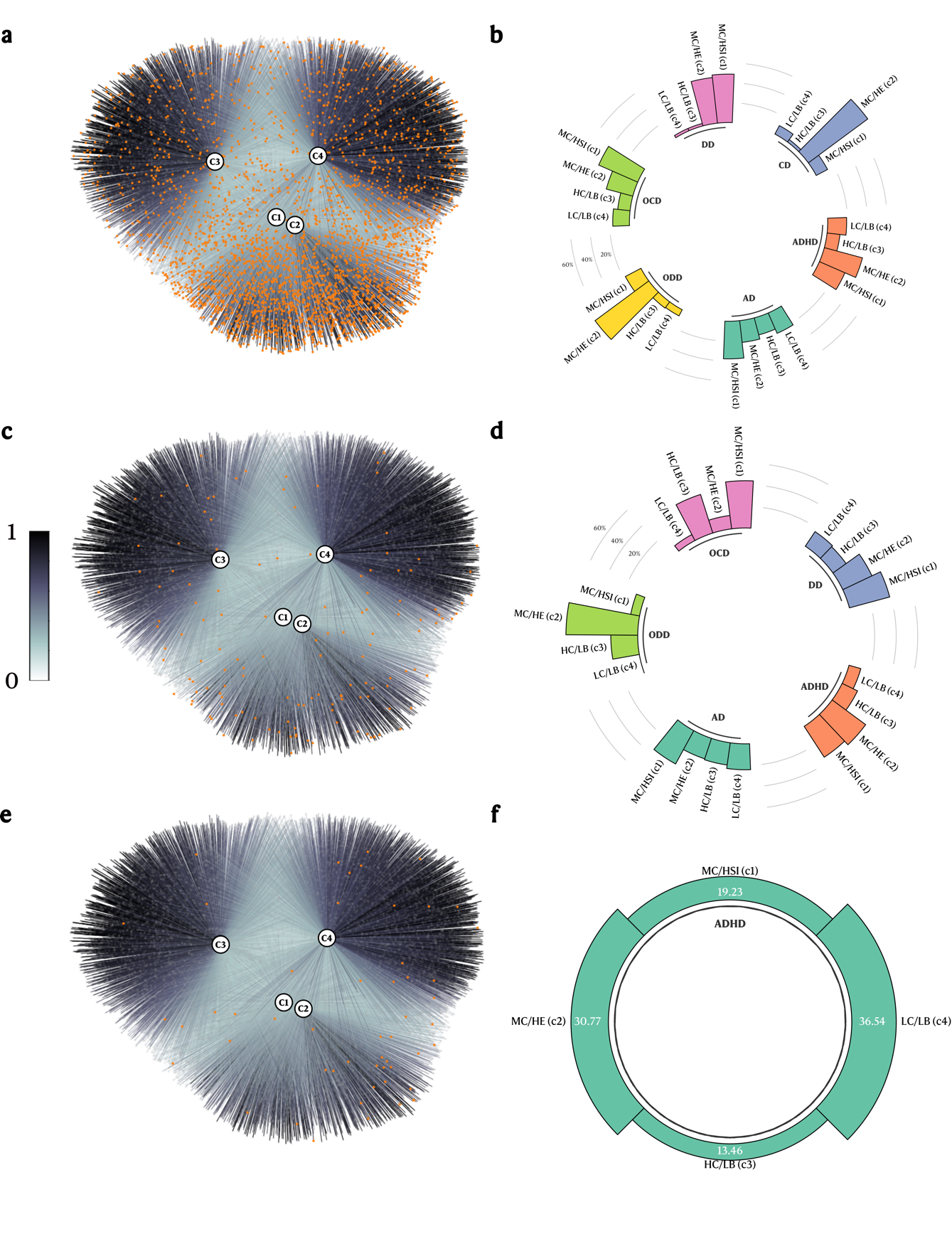

**Supplementary Figure 9.** Diagnosis distribution across all profiles for each independent cohort. **A-B.** ABCD study. **C-D.** BANDA study. **E-F.** GESTE study. Orange node indicates a participant with at least one psychiatric diagnosis. AD: Anxiety Disorder, ADHD: Attention Deficit-Hyperactivity Disorder, CD: Conduct Disorder, DD: Depressive Disorder, OCD: Obsessive-Compulsive Disorder, ODD: Oppositional Defiant Disorder.

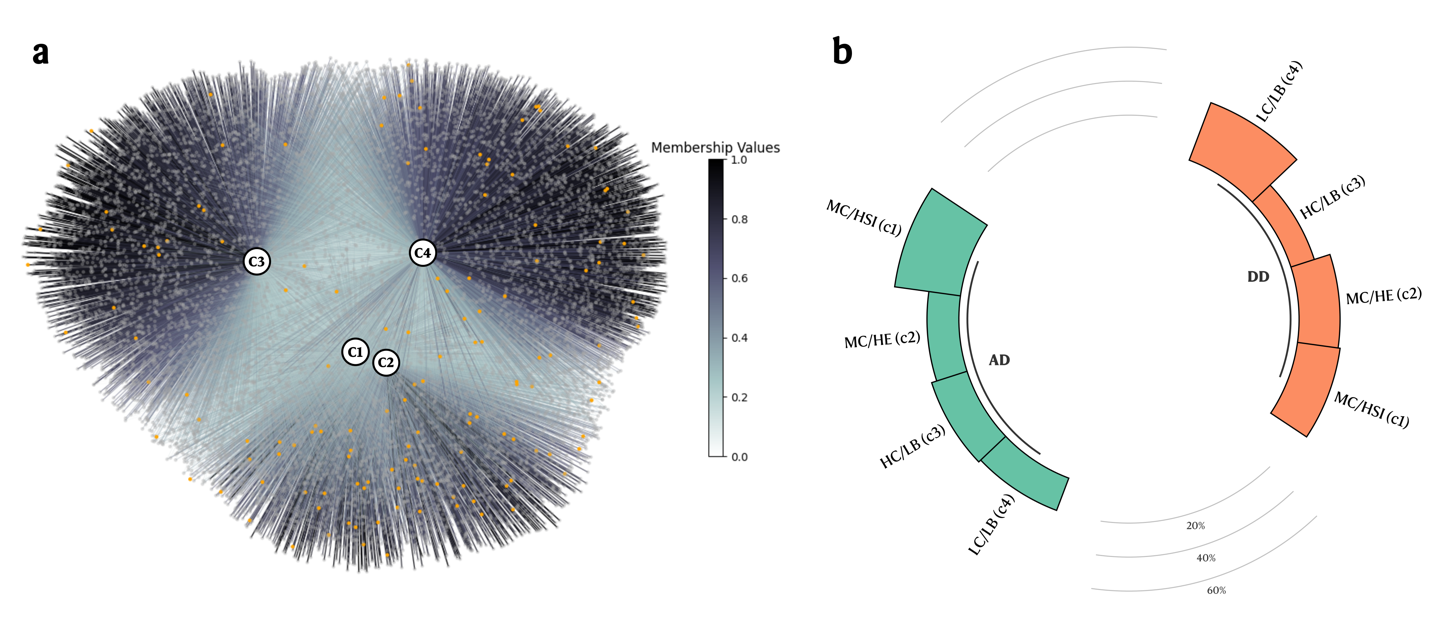

**Supplementary Figure 10.** Youth KSADS diagnosis distribution across the ABCD baseline profiles. **A.** Graph Network with subjects with at least one psychiatric disorder highlighted. **B.** Circular bar plot of the distribution of diagnoses across profiles. AD: Anxiety Disorder. DD: Depressive Disorder.
